# The duration, dynamics and determinants of SARS-CoV-2 antibody responses in individual healthcare workers

**DOI:** 10.1101/2020.11.02.20224824

**Authors:** Sheila F Lumley, Jia Wei, Denise O’Donnell, Nicole E Stoesser, Philippa C Matthews, Alison Howarth, Stephanie B Hatch, Brian D Marsden, Stuart Cox, Tim James, Liam Peck, Thomas Ritter, Zoe de Toledo, Richard J Cornall, E Yvonne Jones, David I Stuart, Gavin Screaton, Daniel Ebner, Sarah Hoosdally, Derrick W Crook, Oxford University Hospitals Staff Testing Group, Christopher P Conlon, Koen B Pouwels, A Sarah Walker, Tim EA Peto, Timothy M Walker, Katie Jeffery, David W Eyre

## Abstract

**Background:** SARS-CoV-2 IgG antibody measurements can be used to estimate the proportion of a population exposed or infected and may be informative about the risk of future infection. Previous estimates of the duration of antibody responses vary.

**Methods:** We present 6 months of data from a longitudinal seroprevalence study of 3217 UK healthcare workers (HCWs). Serial measurements of IgG antibodies to SARS-CoV-2 nucleocapsid were obtained. Bayesian mixed linear models were used to investigate antibody waning and associations with age, gender, ethnicity, previous symptoms and PCR results.

**Results:** In this cohort of working age HCWs, antibody levels rose to a peak at 24 (95% credibility interval, CrI 19-31) days post-first positive PCR test, before beginning to fall. Considering 452 IgG seropositive HCWs over a median of 121 days (maximum 171 days) from their maximum positive IgG titre, the mean estimated antibody half-life was 85 (95%CrI, 81-90) days. The estimated mean time to loss of a positive antibody result was 137 (95%CrI 127-148) days. We observed variation between individuals; higher maximum observed IgG titres were associated with longer estimated antibody half-lives. Increasing age, Asian ethnicity and prior self-reported symptoms were independently associated with higher maximum antibody levels, and increasing age and a positive PCR test undertaken for symptoms with longer antibody half-lives.

**Conclusion:** IgG antibody levels to SARS-CoV-2 nucleocapsid wane within months, and faster in younger adults and those without symptoms. Ongoing longitudinal studies are required to track the long-term duration of antibody levels and their association with immunity to SARS-CoV-2 reinfection.

**Summary:** Serially measured SARS-CoV-2 anti-nucleocapsid IgG titres from 452 seropositive healthcare workers demonstrate levels fall by half in 85 days. From a peak result, detectable antibodies last a mean 137 days. Levels fall faster in younger adults and following asymptomatic infection.

## Introduction

Measurable IgG antibodies to SARS-CoV-2 antigens develop after many, but not all, SARS-CoV-2 infections.[1–4] Serological responses are typically detectable within 1-3 weeks.[5–8] This allows antibody assays to be used to estimate the proportion of a population exposed or infected. Additionally, although the extent of immunity associated with different antibody titres and other immune responses is yet to be fully determined, it is probable that antibody levels will provide some information about the risk and/or severity of future infection.

However, SARS-CoV-2 IgG antibody levels are dynamic over time.[9] This has implications for epidemiological studies, e.g. if IgG levels fall below detection thresholds before they are measured, past infections may be under ascertained. Similarly, if antibodies are a marker for protective immunity, their rate of fall would be of interest when estimating population protection.

Contrasting data have been made available on the longitudinal trajectory and longevity of antibodies induced by SARS-CoV-2 infection. For example, a US study showed IgG antibody levels to trimerised spike were relatively stable in 121 individuals around 110 days post symptom onset.[10] Similarly, data from 1215 individuals in Iceland suggest IgG responses to nucleocapsid and the S1 component of spike were sustained for 100-125 days.[11] However, others have noted declines in neutralizing antibodies over similar time periods.[12–14]

We have recently undertaken baseline serological testing in a cohort of >10,000 HCWs.[15] We now describe quantitative serial SARS-CoV-2 antibody measurements, demonstrating how responses fall over time and vary with age, ethnicity and previous symptoms.

## Methods

### Setting and participants

Oxford University Hospitals NHS Foundation Trust (OUH) offers both symptomatic and asymptomatic SARS-CoV-2 testing programmes to staff at its four teaching hospitals in Oxfordshire, UK. 12,411 healthcare workers (HCWs) have undergone serological testing to date; this analysis includes all 3217 HCWs who attended more than once for antibody testing.

SARS-CoV-2 PCR testing of nasal and oropharyngeal swabs for all symptomatic (new persistent cough, fever ≥37.8°C, anosmia/ageusia) staff was offered from 27-March-2020 onwards. Asymptomatic HCWs were invited to participate in voluntary staff testing for SARS-CoV-2 by nasal and oropharyngeal swab PCR and serological testing from 23-April-2020 onwards. The cohort, associated methods and findings from the first test per individual have been previously described.[15] Following initial PCR and antibody testing, asymptomatic HCWs were invited to optionally attend for serological testing up to once every two months, with some offered more frequent screening as part of related studies. Asymptomatic staff were also offered optional SARS-CoV-2 PCR tests every two weeks. The symptomatic programme continued to offer on-site PCR testing whenever a staff member developed symptoms.

### Laboratory assays

Serology for SARS-CoV-2 IgG to nucleocapsid protein was performed using the Abbott Architect i2000 chemiluminescent microparticle immunoassay (CMIA; Abbott, Maidenhead, UK). Antibody levels ≥1.40 manufacturer’s arbitrary units were considered positive, 0.50-1.39 equivocal (following Abbott Diagnostics Product Information Letter PI1060-2020) and <0.5 negative.

### Statistical methods

Individuals with ≥1 positive antibody result (titre ≥1.40) and ≥2 antibody results were classified as showing rising titres only, falling or stable titres only, or both. Those with only one measurement could not be classified and were excluded. In those with falling/stable titres we estimated the duration of antibody responses from the maximum observed result using Bayesian linear mixed models and their association with age, gender, ethnicity, previous self-reported symptoms and PCR results (allowing correlated random intercept and slope terms, see Supplement). We assumed antibody levels fell exponentially, and so modelled log2 transformed antibody levels over time (observed data and fitted models demonstrated close congruence, Supplementary File). The incidence of Covid-19 in our hospital fell after a peak in March and April 2020,[15] such that re-exposure of HCWs was uncommon; we therefore had insufficient data to study boosting of antibody responses.

We additionally modelled the antibody trajectory from a first positive PCR test using a similar approach but allowing for non-linear effects of time rather than assuming an exponential decline.

We used Bayesian interval censored regression to estimate the proportion of individuals remaining antibody positive at varying times following their maximum antibody result (see Supplement).

### Ethics

Deidentified data from staff testing were obtained from the Infections in Oxfordshire Research Database (IORD) which has generic Research Ethics Committee, Health Research Authority and Confidentiality Advisory Group approvals (19/SC/0403, 19/CAG/0144).

## Results

A total of 3217 HCWs provided ≥2 samples for serological testing between 23-April and 20-October-2020 (Figure 1, Table 1). The median (IQR) [range] number of samples tested was 2 (2-2) [2-8] and time from the first to last sample was 124 (95-144) [3-174] days.

**Table 1.** Baseline cohort demographics for 3217 HCWs and 452 HCWs with ≥1 positive SARS-CoV-2 IgG result and ≥1 subsequent follow up sample.

| Characteristic | Whole cohort<br>n (%) or median (IQR) [Range] | 452 HCWs with a positive<br>antibody result and ≥1<br>subsequent sample<br>n (%) or median (IQR) [Range] |
| --- | --- | --- |
| **Age** |  |  |
| Age, years | 39 (29-50) [16-76] | 41 (29-50) [17-69] |
| **Gender** |  |  |
| Female | 2542 (79) | 340 (75) |
| Male | 673 (21) | 112 (25) |
| Not disclosed | 2 (<1) |  |
| **Self-reported ethnicity** |  |  |
| White | 2473 (77) | 302 (67) |
| Black | 90 (3) | 25 (6) |
| Asian | 440 (14) | 89 (20) |
| Other | 214 (7) | 36 (8) |
| **Covid-19-like symptoms between 01**<br>**February 2020 and testing** |  |  |
| Yes | 898 (28) | 274 (61) |
| No | 2319 (72) | 178 (39) |
| **Previous positive SARS-Cov-2 PCR** |  |  |
| Symptomatic | 128 (4) | 95 (21) |
| Asymptomatic | 117 (4) | 59 (13) |
| None | 2972 (92) | 298 (66) |

**Figure 1.**
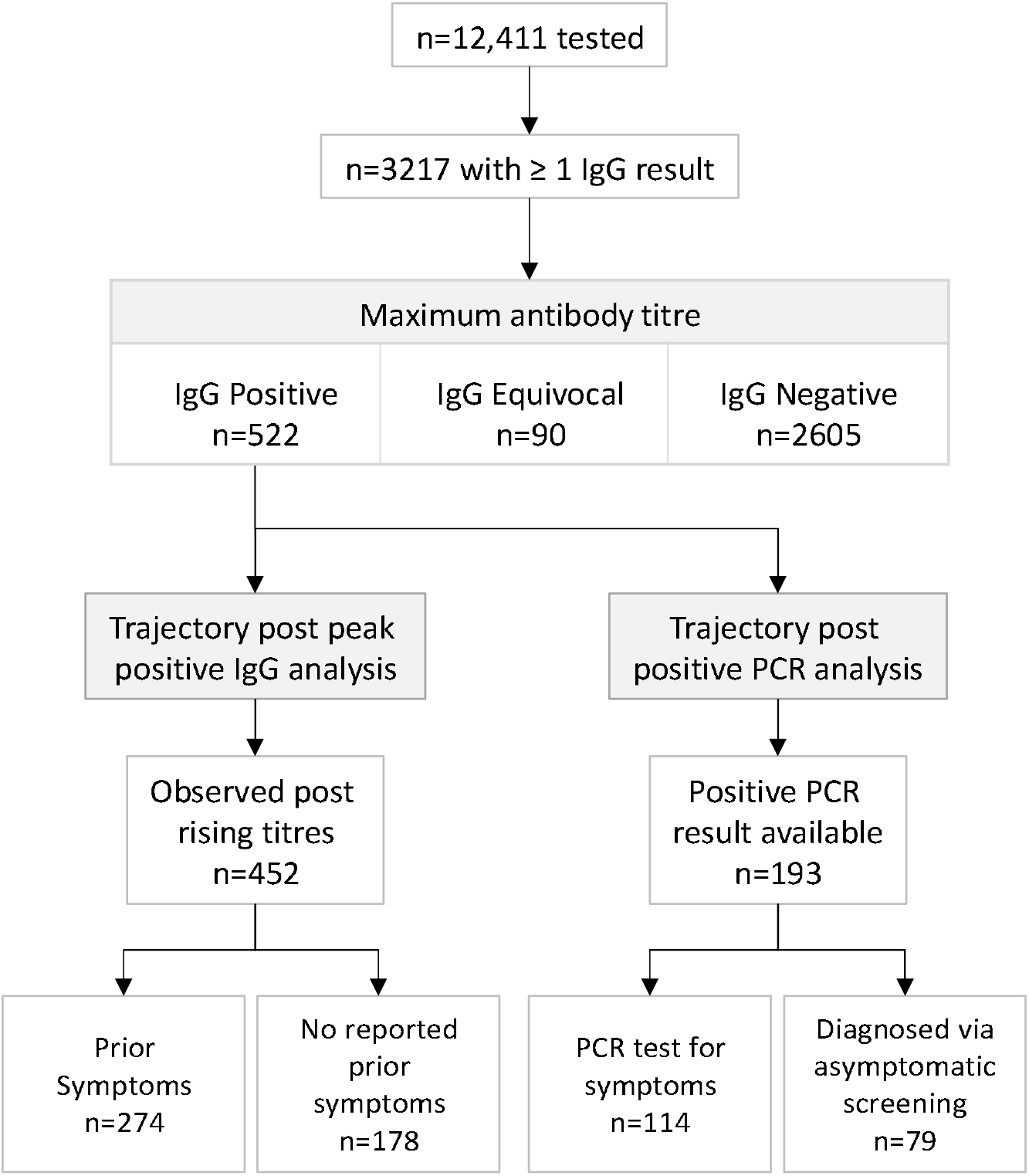
SARS-CoV-2 antibody trajectory cohorts.

Observed IgG antibody trajectories are shown in Figure 2. 522 (16%) HCWs had ≥1 sample with detected SARS-CoV-2 IgG antibodies (titre ≥1.40; Figure 2A, 2B) and 90 (3%) had ≥1 sample with an equivocal maximum IgG titre (0.50-1.39; Figure 2C). Antibody titres in those with consistently negative results were broadly stable (Figure 2D), whereas falls were observed in 438/466 (94%) initially positive individuals, and in 61/83 (73%) with an initial equivocal titre (Figures 2A, 2C, 3). Consistent with prior infection in some individuals with equivocal titres and subsequent waning, there were more falls in antibody levels than expected by chance in those with titres of 0.80-1.39 (Figure 3).

**Figure 2.**
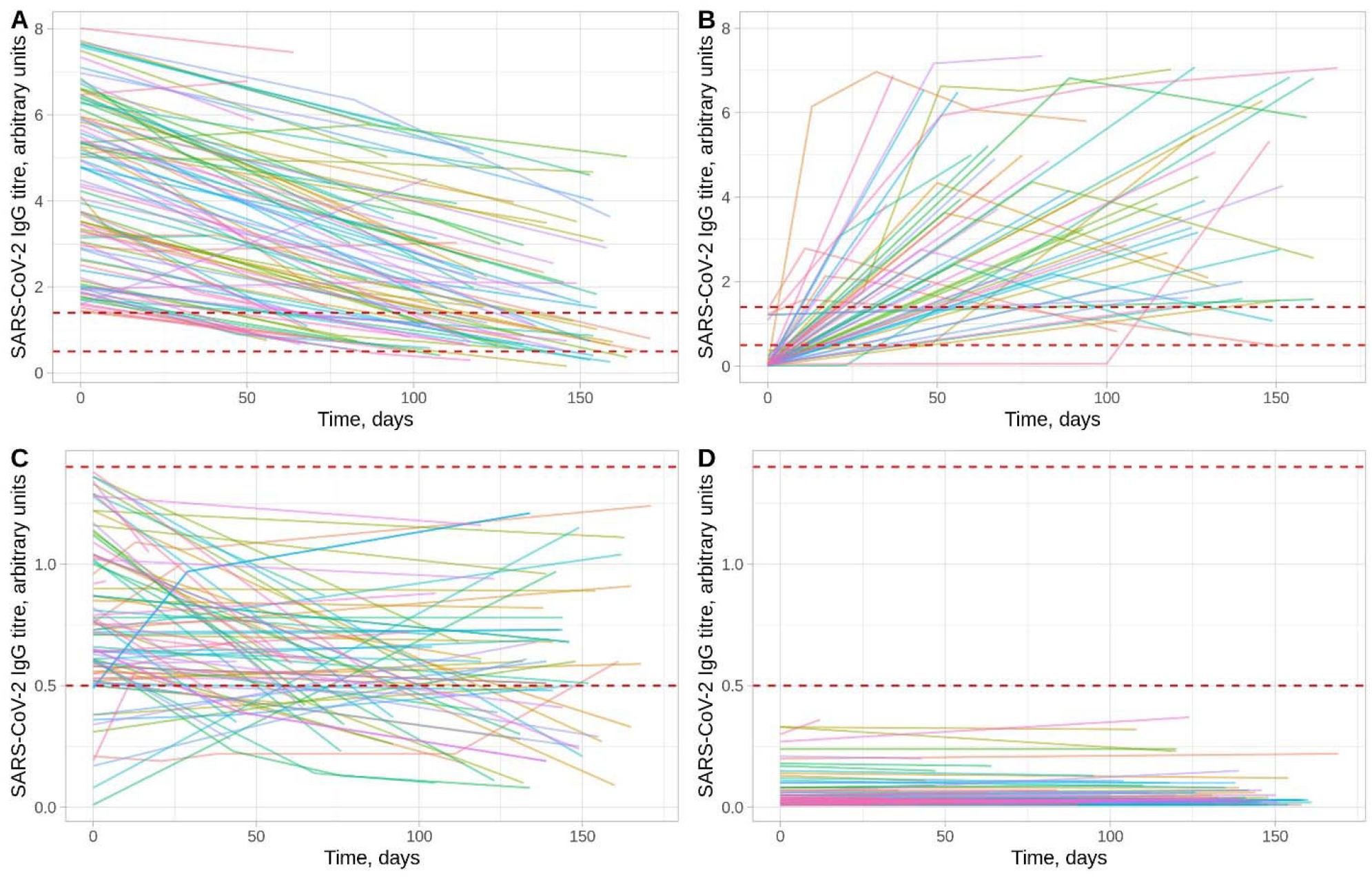
SARS-CoV-2 antibody trajectories in 3217 HCWs. Panels A and B show trajectories for HCWs with a positive result (≥1.40 arbitrary units) at some time. Panel A shows those whose first measurement was positive (n=466, only data from 100 randomly sampled individuals is shown to assist visualisation) and Panel B the remainder (n=56) in whom seroconversion was observed. Panel C shows those with a maximum titre that was equivocal (0.50-1.39, n=90). Panel D shows results from HCWs with a maximum titre that was negative (<0.50, n=2605, only data from 100 randomly sampled individuals is shown to assist visualisation). The dashed lines indicate the thresholds for a positive and equivocal result, note the different y-axis scales in panels A and B versus panels C and D.

**Figure 3.**
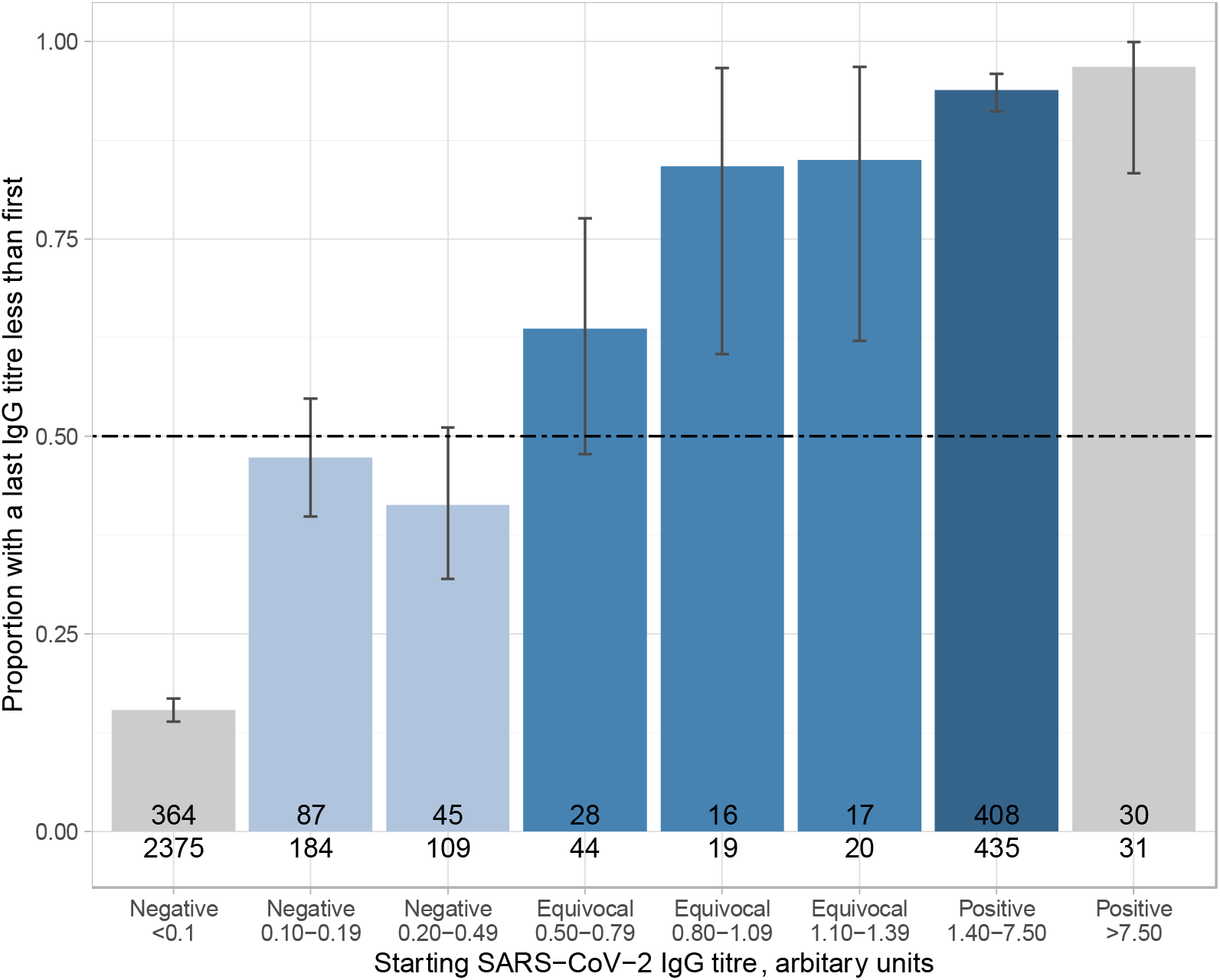
Proportion of 3217 HCWs with an observed fall in antibody titres over time. Titres <0.1 and >7.50 are shown in grey to indicate that the initial measurements are near the lower and upper limits of quantification and so the proportion observed for these bars is likely to be biased and should be discounted. Light, mid and dark blue bars represent initially negative, equivocal and positive antibody titres respectively. Under a random model the observed proportion of individuals with a last IgG titre less than the first is expected to be 0.50 as shown by the dashed line. The numerator and denominator for each bar is shown. Most individuals were sampled for the first time from May 2020 onwards, i.e. several weeks after the local peak in the first wave of the pandemic, such that an excess of falling titres is expected to consistent with previous infection.

### Antibody trajectories after a positive IgG test

Among 522 individuals with ≥1 IgG-positive sample, 70 (13%) showed evidence of seroconversion with rising titres only and so were excluded from analyses of the duration of response following a positive IgG result (39/70 were PCR-positive and are included in a separate analysis below of responses following a positive PCR test). In the remaining 452 (87%), the median (IQR) [range] number of samples tested was 2 (2-3) [2-5] and time from the first to last sample was 121 (83-143) [4-171] days. Only 3/120 (3%) individuals with ≥3 measurements had a final titre above the minimum observed, i.e. potential evidence of boosting, and titre increases were small (6.30→6.59, 2.48→2.60, 1.22→1.25). The median (IQR) age was 41 (29-50) years and 75% of participants were female (Table 1). The most common self-reported ethnic groups were White (302, 67%) and Asian (89, 20%; predominately south Asian and Filipino). 274 (61%) recalled self-identified Covid-19-like symptoms between 01-February-2020 and testing. 95 (21%) had a positive SARS-CoV-2 PCR following symptomatic testing and 59 (13%) a positive PCR during asymptomatic screening. It is likely that many of the remainder were infected prior to widespread availability of symptomatic testing or asymptomatic screening. The first positive PCR in each individual was prior to or on the day of their maximum antibody titre in all but 5/154 (3%) (who were tested 3, 8, 8, 11 and 17 days later).

Using a Bayesian statistical model, the trajectory of SARS-CoV-2 IgG antibody levels following the maximum measured antibody titre in each individual is shown in Figure 4A. The estimated mean antibody half-life was 85 (95% credibility interval, CrI 81-90) days. The estimated mean maximum antibody level was 4.3 (95%CrI 4.1-4.4) arbitrary units. The mean trajectory crossed the diagnostic threshold of 1.40, switching from a positive to equivocal result at 137 (95%CrI 127-148) days. IgG level half-lives and starting maximum titres varied between individuals (Figures 4B and 4C respectively). Higher maximum observed IgG levels were correlated with longer estimated IgG half-lives, i.e. slower rates of decline over time (Figure 4D; Spearman’s rank R^2^=0.65, p<0.0001). Considering only maximum levels ≤7.50 to avoid titres near the upper limit of quantification yielded similar results (R^2^=0.61, p<0.0001).

**Figure 4.**
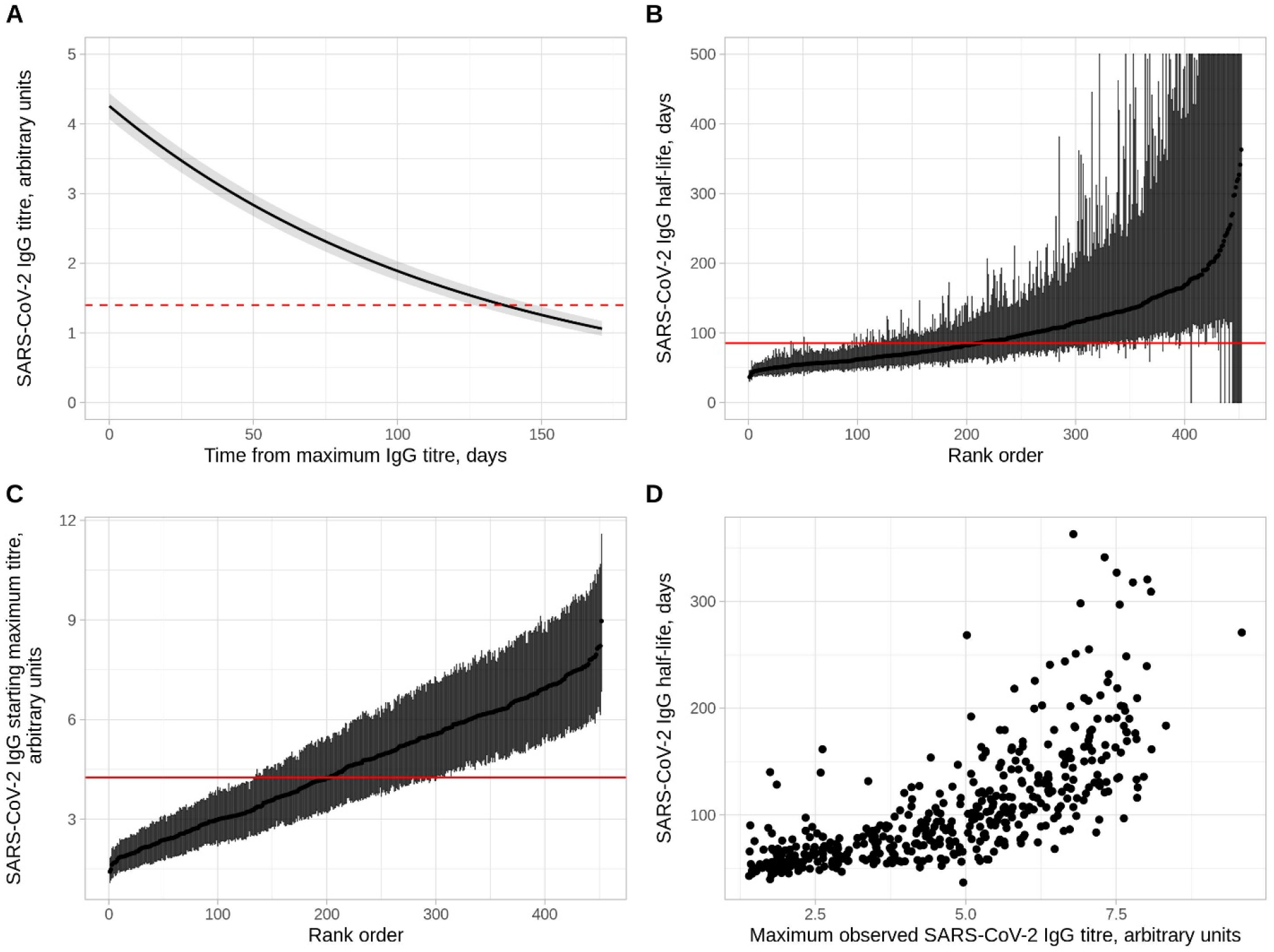
Anti-nucleocapsid IgG antibody trajectories in 452 SARS-CoV-2 seropositive HCWs. Panel A shows the overall mean trajectory of SARS-CoV-2 IgG antibody levels from the maximum observed level (i.e. the model fixed effect). The posterior mean and 95% credibility interval are shown as a solid line and shaded area. IgG antibody levels are shown in arbitrary units from the Abbott CMIA. The dashed red line represents the diagnostic threshold of 1.40 arbitrary units. Panel B shows the estimated SARS-CoV-2 IgG half-life with 95% CrI by days for all participants, ranked by its value. The solid red line indicates the overall mean. Credibility intervals exceeding 500 days are truncated at 500 days. Panel C shows the estimated maximum antibody level with 95% CrI for all participants, ranked by its value. The solid red line indicates the overall mean. Panel D shows a comparison of maximum observed SARS-CoV-2 IgG antibody level and the estimated SARS-CoV-2 IgG half-life per individual.

To facilitate comparison with studies reporting only categorical antibody results, we considered the proportion of seropositive individuals remaining antibody-positive (as opposed to equivocal or negative) when observed at varying time intervals (Figure 5A) and using an interval censored survival analysis approach (Figure 5B). Consistent with the model in Figure 4, the median time remaining antibody positive was 166 (95%CrI 139-214) days.

**Figure 5.**
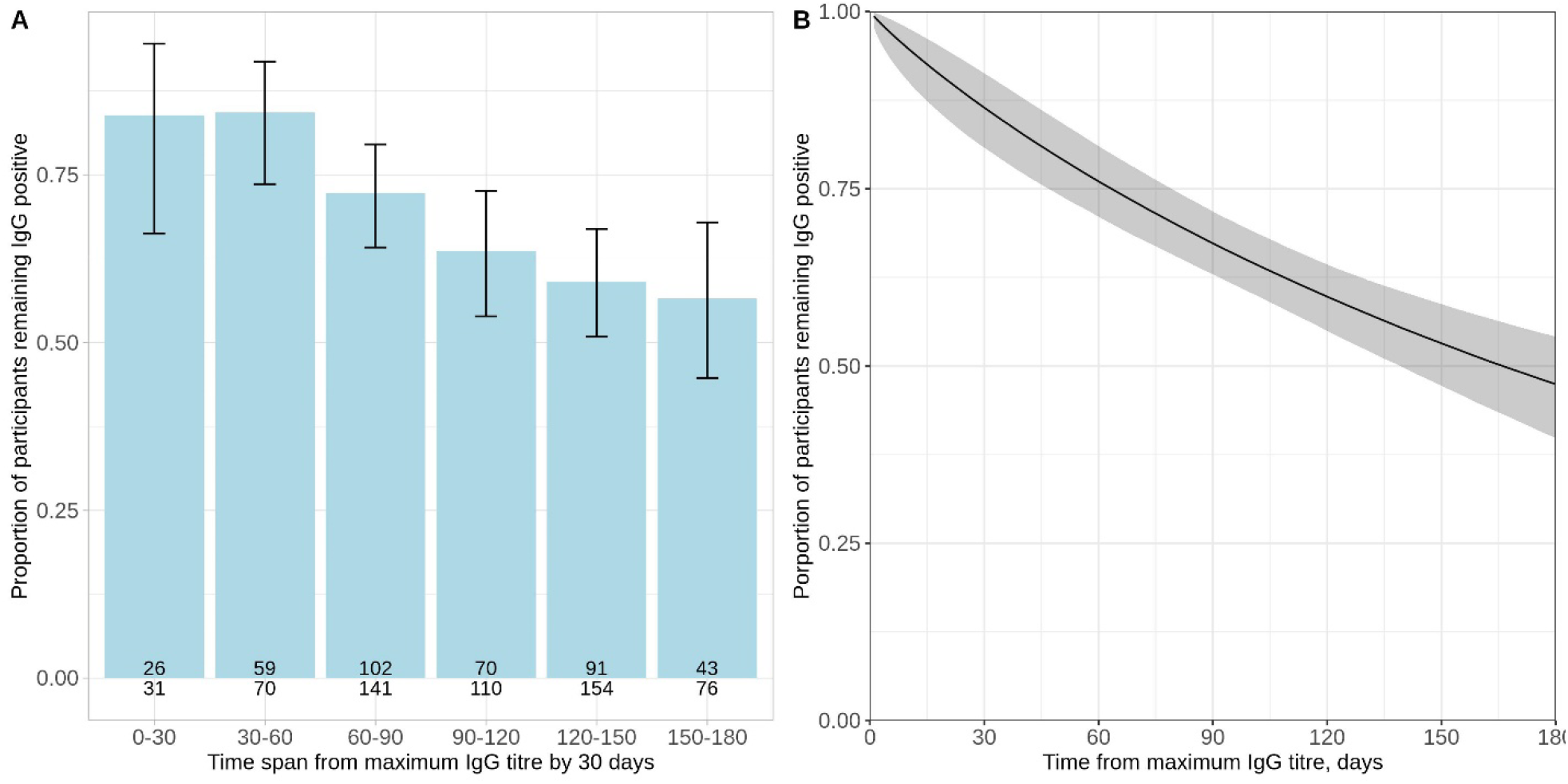
Proportion of HCWs remaining antibody positive by days following their maximum antibody level. Panel A shows the observed proportion in 30-day intervals with binomial 95% confidence intervals. The number of individuals tested and the number of individuals remaining antibody positive is shown at the base of each bar. Panel B shows the results of a Bayesian interval censored survival analysis, the posterior mean and 95% credibility interval are shown.

### Effect of demographics and other covariates

Age, self-reported ethnicity, prior symptoms compatible with Covid-19 and a positive SARS-CoV-2 PCR were independently associated with changes in antibody trajectories: affecting the starting maximum level (the intercept), the antibody half-life (the slope) or both (Table 2, Figure S1, details in Figures S2-S4 and Table S2).

**Table 2.**
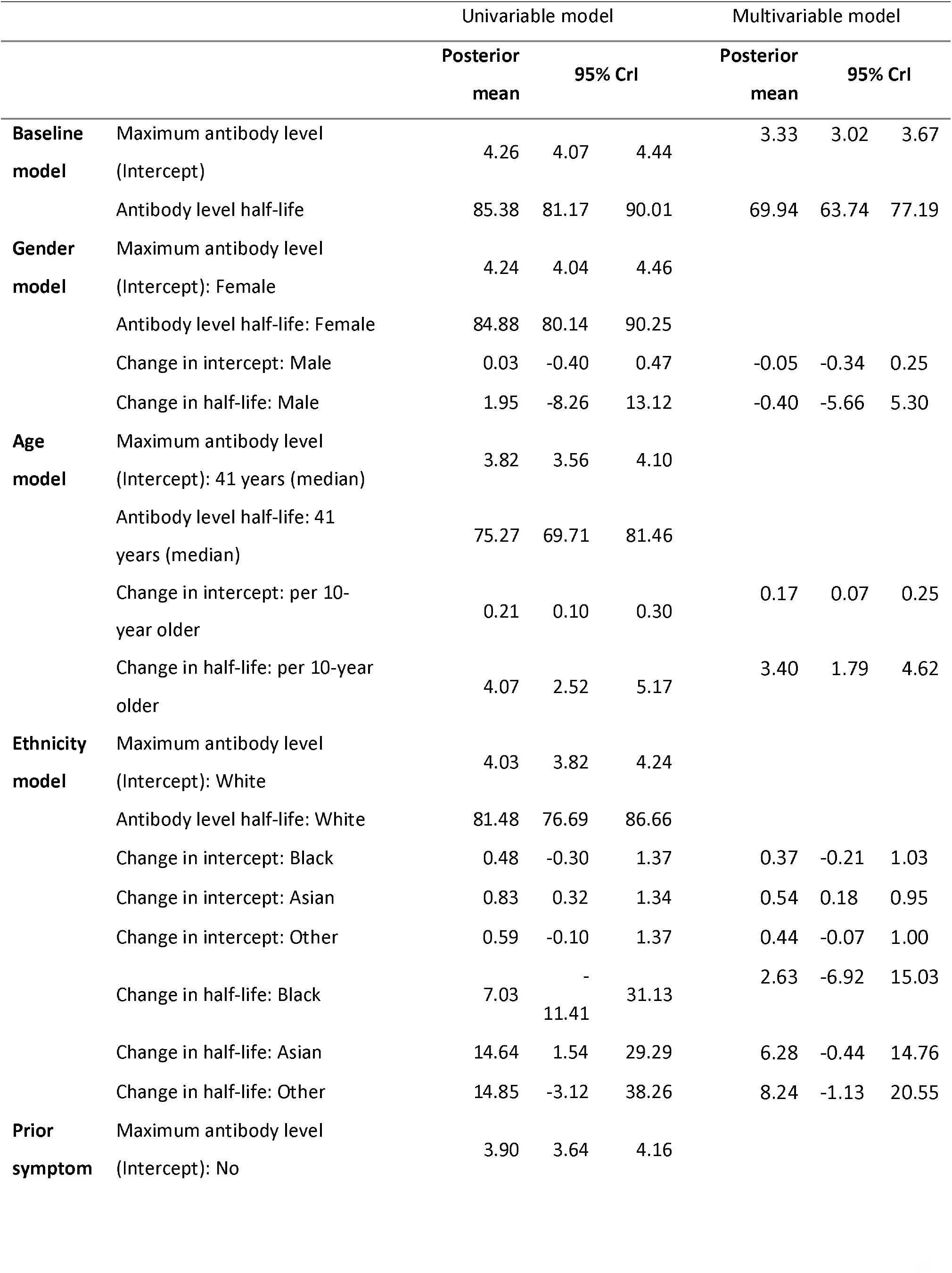
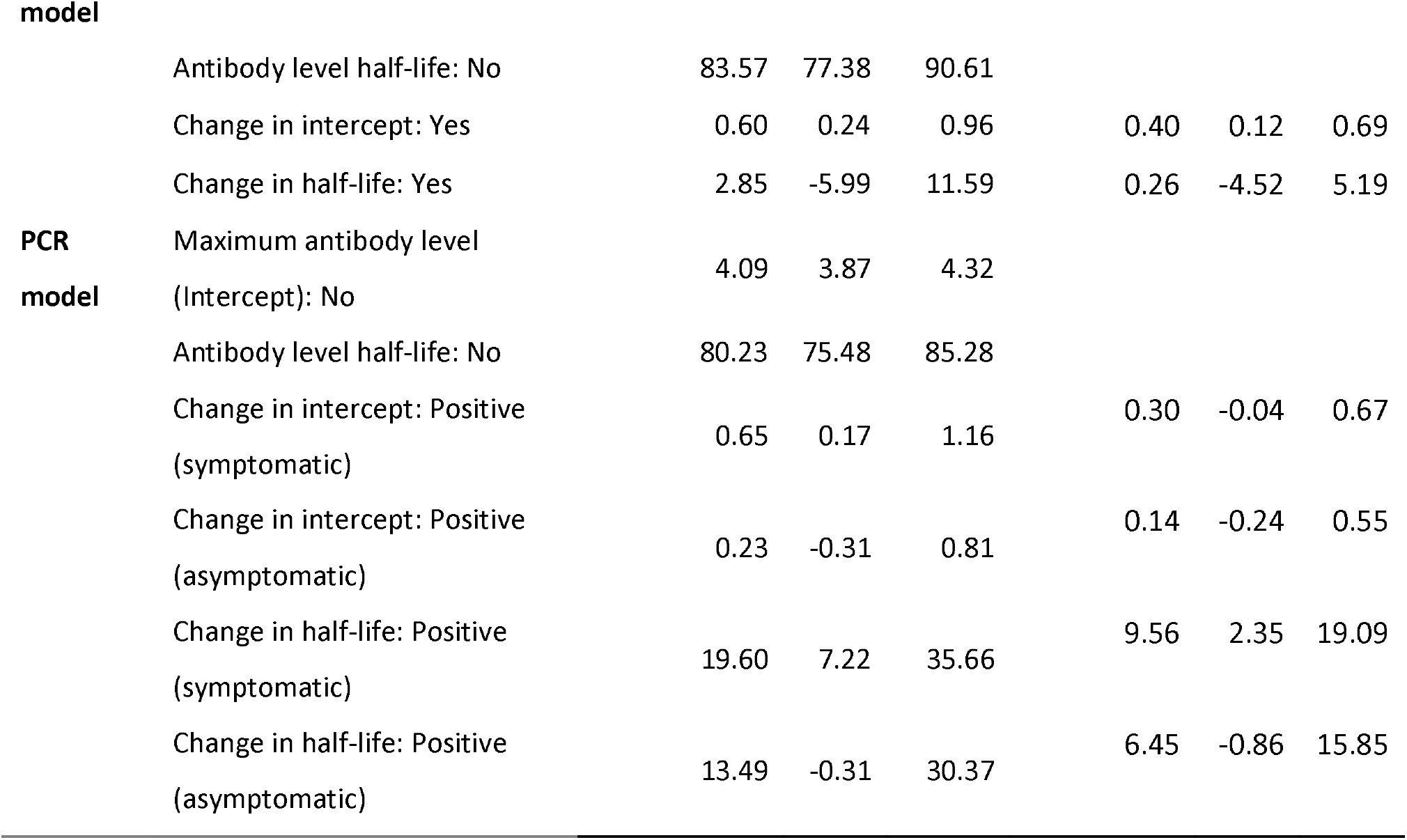
Univariable and multivariable models of determinants of SARS-CoV-2 antibody trajectories. Posterior mean and 95% credibility intervals for the maximum antibody level (model intercept) and antibody half-life (model slope) are shown. See Supplementary Table S2 for other model parameters and statistical analysis quality metrics.

Within this cohort of HCWs of working age, increasing age was independently associated with higher maximum antibody levels and a longer IgG half-life, 0.17 (95%CrI 0.07-0.25) arbitrary units and 3.40 (95%CrI 1.79-4.62) days per 10 years respectively (Table 2, Figure S5).

HCWs of Asian ethnicity had higher maximum antibody levels with adjusted increases of 0.54 (95%CrI 0.18-0.95) arbitrary units compared to White HCWs, with marginal evidence for longer antibody half-lives, by 6.28 (−0.44 to 14.8) days. Within the limits of the power of the study, there was no strong statistical evidence that antibody trajectories varied in other ethnic groups.

Prior self-reported symptoms were associated with a higher starting maximum level (adjusted increase 0.40 [95%CrI 0.12-0.69]), but not changes in antibody half-lives. There was moderate evidence that a positive PCR result arising from testing undertaken for symptoms, independently increased the starting maximum level by 0.30 (95%CrI -0.04 to 0.67) arbitrary units, and half-life by 9.56 (95%CrI 2.35-19.09) days compared to those with no positive PCR. We observed no effect of gender with either maximum level or antibody half-life.

### Sensitivity analysis

109 participants had ≥2 IgG readings after their maximum observed antibody titre and were included in a sensitivity analysis to investigate the impact of starting with each individual’s maximum result on half-life estimates (which should be independent of starting values under the exponential decay assumption). Using only observations after the maximum antibody titre, the estimated half-life was 83.0 (95%CrI 71.8-97.2) days, consistent with the main analysis, with a lower estimated intercept (2.8 [95%CrI 2.5-3.1]), as expected.

### Antibody trajectories following a positive PCR test

245 of the 3217 HCWs with ≥2 serology samples had a positive PCR test. 114/128 (89%) symptomatic HCW seroconverted (maximum IgG titre ≥1.40), including 11/12 (92%) who required hospital treatment, all other infections were mild. 79/117 (68%) identified through asymptomatic screening seroconverted. In the 52 individuals not showing evidence of seroconversion, all but 1 (98%) had ≥1 antibody test ≥14 days after their PCR-positive test, and in 29 (56%) this was before 90 days. PCR cycle threshold values were lower in individuals who seroconverted (Table S3).

Data from PCR-positive individuals who seroconverted were used to model antibody trajectories relative to a first positive PCR test. Antibody levels rose to a peak at 24 (95%CrI 19-31) days post-first positive PCR test, before beginning to fall (Figure 6). Comparing with the antibody trajectory estimated in the main analysis, the estimated rates of waning were consistent between the two models. Antibody trajectories were similar in those being tested following symptoms or during asymptomatic screening (Figure S6).

**Figure 6.**
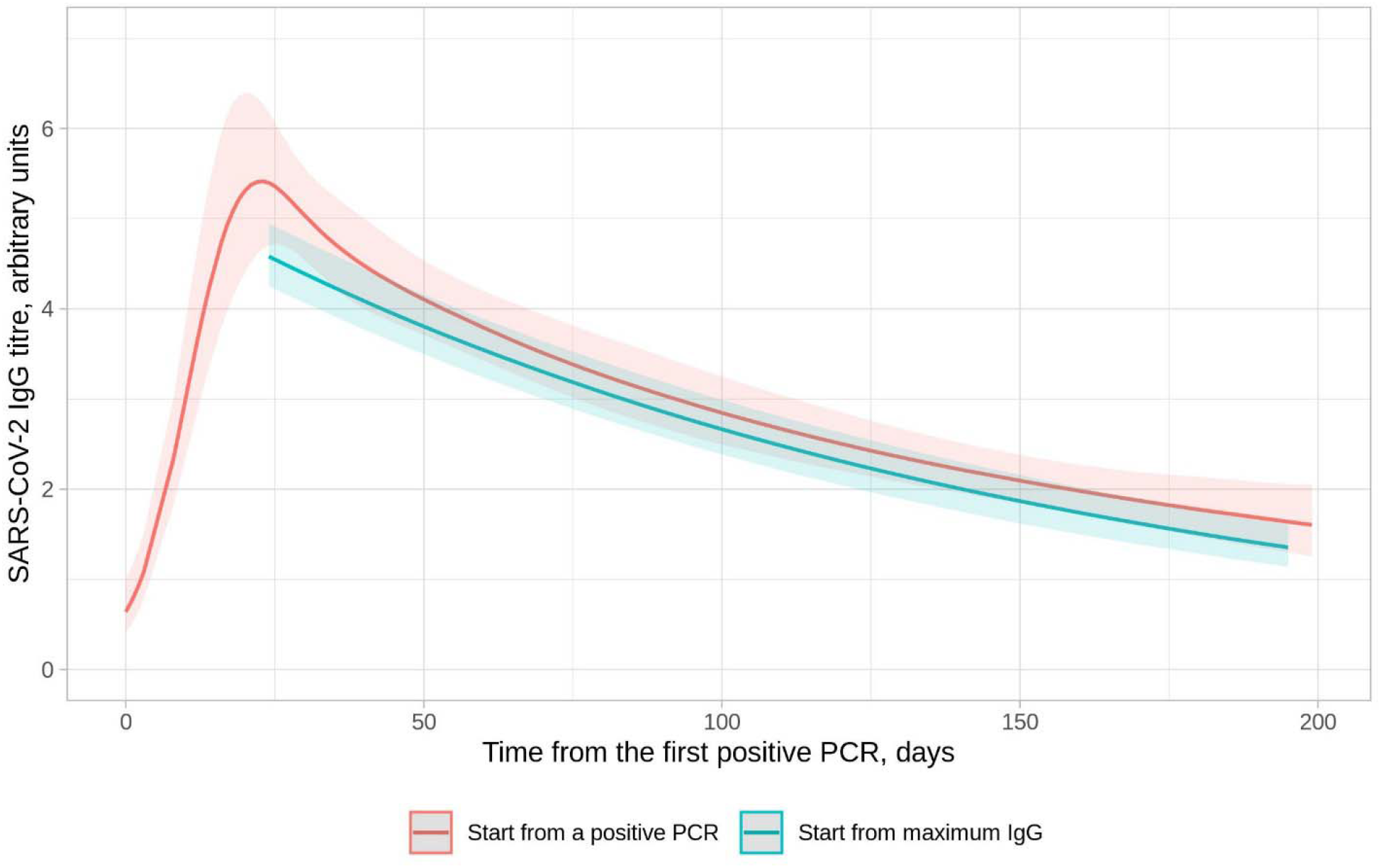
Comparison of SARS-CoV-2 IgG antibody levels following a positive PCR test and the maximum IgG level per individual in those with a positive PCR test. The x-axis value for the model starting from the maximum IgG level is aligned to the maximum point from the model starting with a positive PCR test. The model starting from a positive PCR is fitted with a 5-knot spline (3 interior knots at t=10, t=30, and t=50, locations chosen based on model fit).

## Discussion

Most epidemiological outbreak models assume that SARS-CoV-2 infection leads to the development of post-infection immunity for a defined duration. Here we describe the antibody trajectories of 452 SARS-CoV-2 IgG-seropositive HCWs over a median 121 days of follow-up from their maximum IgG titre. By following quantitative antibody responses, we could separately analyse changes in initial antibody levels and rates of waning. The estimated antibody half-life was 85 (95%CrI 81-90) days. We observed variation between individuals; higher maximum observed IgG titres were associated with longer half-lives. Increasing age, Asian ethnicity and prior self-reported symptoms were independently associated with higher maximum antibody levels, and increasing age and a positive PCR test undertaken for symptoms with longer antibody half-lives. 39% of HCWs were asymptomatic throughout, and therefore our data also represent an important contribution to the literature, which to date has mainly focused on trajectories following symptomatic infection.

The durability of SARS-CoV-2 IgG seen here is more similar to the seasonal coronaviruses, where IgG waning and reinfection can occur within a year,[16] rather than to other epidemic coronaviruses, SARS-CoV and MERS-CoV, where most long-term studies have found that although IgG wanes over time, it is detectable up to 1-3 years later.[17] However, to some extent these findings are conditional on assay cut-offs which can be tuned to prioritise sensitivity or specificity. We observe higher IgG titres with longer durability occurring after symptomatic PCR-positive infection, consistent with data from Long et al. where 40% of asymptomatic individuals and 13% of the symptomatic group became negative for IgG in the early convalescent phase[18] and consistent with emerging coronaviruses, where antibody titres remained detectable longer after more severe illness,[17] waning more rapidly after asymptomatic infection.

Relatively short-term antibody responses have two epidemiological consequences. Firstly, antibody waning may lead to under-ascertainment of previous infections within the current pandemic, particularly in younger individuals following asymptomatic/mild infection. Additionally, IgG testing is unlikely to determine whether SARS-CoV-2 has circulated historically, e.g. in a particular geographic region. Older age (within this cohort of working age HCWs, up to 69 years) was associated with higher maximum observed IgG titres and longer half-lives. It is possible to hypothesise that this could arise from boosting of cross-reactive antibodies from prior exposure to a previously circulating human coronavirus. However, at least in the absence of recent boosting, circulating cross-reactive antibodies to SARS-CoV-2 spike are uncommon and cross-reacting neutralizing antibodies even rarer.[19] Furthermore, although there is some cross-reactivity between seasonal coronaviruses in the same genetic group after natural and experimental infection, there is little cross-reactivity between seasonal coronaviruses and the emerging coronaviruses SARS-CoV and MERS-CoV.[17,20] We also see higher maximum observed IgG titres with moderate evidence of longer half-lives in Asian HCWs. Many of these individuals are from south Asia and the Philippines and were recruited overseas as adults to work in the UK healthcare system. It is possible to speculate that higher levels in this group may reflect geographically-restricted exposures to similar viruses in the past, but for the reasons outlined above, the evidence for this is not strong.

### Limitations

One limitation of our study was that our cohort of individuals consisted of adults of working age (17-69 years); further longitudinal studies will be required to investigate antibody trajectories in younger and older age groups. In addition, the small numbers of self-reported Black (n=25) and Other (n=36) ethnicities reduced/limited power to detect an association between these ethnicities and antibody trajectories. Furthermore, we do not account for multiple mediators, e.g. socioeconomic inequalities, that may link ethnicity to antibody responses in the absence of a direct causal relationship. Due to many of our staff developing symptoms before widespread SARS-CoV-2 PCR testing was available, only 34% of the antibody positive cohort had a documented positive PCR, and as a proportion of the cohort were asymptomatic throughout, we modelled time from maximum positive antibody test rather than time from first positive PCR or time from symptom onset in our main analysis of antibody durability. However under an exponential assumption, half-lives can be unbiasedly estimated from any measurements taken after a maximum; we excluded individuals with only evidence of rising titres to avoid underestimating half-lives. Further, data from those that were PCR positive were consistent with this analysis (Figure 6).

Multiple different assays are in use globally to characterize antibody responses to SARS-CoV-2. Here we use a commercially-available anti-nucleocapsid IgG assay; however, other antibody classes and targets, and aspects of immunity, including the innate and cellular responses are important in conferring post-infection immunity.[21] When comparing longitudinal studies of antibody durability, care must be taken, as the various assays have not yet been cross-calibrated, and implications for protective immunity are not fully understood.

### Implications

It is widely recognized that pathogen-specific IgG levels decline after the acute phase of an infection. After the initial humoral response in which short-lived plasmablasts secrete high titres of antibody, long-lived plasma cells and memory B cells then contribute to longer-term antibody-mediated protection.[22] Although declines in IgG titres are expected, understanding the rate of decline, whether and when titres fall below assay positive cut-offs, and how these titres relate to protection from subsequent asymptomatic and symptomatic re-infection is crucial.

Serological testing also helps quantify the extent of infection in populations, informing epidemiological models which direct public health strategies including nonpharmaceutical interventions. Seroprevalence measures, such as IgG antibody titres, are imperfect, as we describe, antibody levels within individuals wane, potentially leading to underestimated exposure due to loss of seropositivity. For example, in this study an estimated 33% of individuals seroreverted (i.e. fell below the positive cut off for the Abbott assay) within 3 months of IgG detection and an estimated 53% by 6 months (Figure 5B). This study highlights that sero-epidemiological surveys performed several months into this pandemic are likely to underestimate prior exposure in younger adults, as they tend to lose detectable antibody faster. Our findings contrast with the repeated cross-sectional REACT2 study,[23] which reported greater reductions over time at a population-level in the proportion of adults ≥65 years testing antibody-positive, and more sustained responses in those 18-24 years. Nearly all HCWs in our study were <65 years, other differences may arise from study design, the assay used, and the potential for new infections predominantly in younger people[24] to replace others who had sero-reverted, supporting population-level seroprevalence (in our study these individuals are followed separately). Ongoing longitudinal studies are required to determine the long-term kinetics of antibody-mediated response to SARS-CoV-2, and responses to re-exposure.

### Future work

Antibody dynamics have significant implications for the course and management of pandemics. Durability of immunity post-infection and post-vaccination will dictate the overall course of the pandemic. Further work is required to determine how prior infection and/or vaccination impacts the probability of future infection and severity of subsequent disease, determine the antibody-based correlates of this protection, and therefore the ability of serological tests to identify those who are immune. Longitudinal cohorts with baseline immunology are required to determine immune correlates of protection, to determine whether measurement of the current antibody status is enough to infer whether an individual have functional immunity or not, whether waning IgG titres are representative of waning immune protection, or whether protection remains even after an individual seroreverts.

## Conclusion

We demonstrate that the half-life of SARS-CoV-2 anti-nucleocapsid IgG antibody responses in a cohort of adult HCWs is 85 days and varies between individuals by age, ethnicity and prior symptom history. The extent and duration of immunity to SARS-CoV-2 infection following Covid-19 and its association with antibody titres remains a key question to be answered.

## Data Availability

The data studied are available from the Infections in Oxfordshire Research Database (https://oxfordbrc.nihr.ac.uk/research-themes-overview/antimicrobial-resistance-and-modernising-microbiology/infections-in-oxfordshire-research-database-iord/), subject to an application and research proposal meeting the ethical and governance requirements of the Database. For further details on how to apply for access to the data and for a research proposal template please.

## Acknowledgements

We would like to thank all OUH staff who participated in the staff testing programme, and the staff and medical students who ran the programme. This work uses data provided by healthcare workers and collected by the UK’s National Health Service as part of their care and support. We thank all the people of Oxfordshire who contribute to the Infections in Oxfordshire Research Database. Research Database Team: L Butcher, H Boseley, C Crichton, DW Crook, DW Eyre, O Freeman, J Gearing (community), R Harrington, K Jeffery, M Landray, A Pal, TEA Peto, TP Quan, J Robinson (community), J Sellors, B Shine, AS Walker, D Waller. Patient and Public Panel: G Blower, C Mancey, P McLoughlin, B Nichols.

