## Supplementary Material for "The duration, dynamics and determinants of SARS-CoV-2 antibody responses in individual healthcare workers"

### Supplementary methods

#### Statistical methods

We used Bayesian linear mixed models to analyse the duration of antibody responses, starting from each individual’s measured maximum antibody level. We assumed antibody levels fell exponentially, and so modelled the log2 transformed antibody level over time. We allowed for correlated random intercept and slope terms to allow for per individual variation. We analysed the univariable effect of covariates of interest on the fixed effect intercept and slope terms, including age, gender, ethnicity, recall of prior symptoms compatible with Covid-19, and a previous positive SARS-CoV-2 PCR test (undertaken either following symptoms or as part of asymptomatic screening). Multivariable models were also fitted, undertaking variable selection based on the leave-one-out cross-validation information criterion (LOOIC), calculated using Pareto smoothed importance sampling, or k-fold cross-validation were Pareto k was >0.7. However, there was no evidence than any model was a better fit than the full multivariable model, and so this is presented. We allowed for non-linear effects of age by using natural cubic splines (R “ns” command) with up to 5 knots using default locations, choosing the best fitting model based on LOOIC.

Our approach is potentially subject to two biases. The first arises from regression to the mean arising from random measurement error, because the analysis is conditional on starting the with maximum antibody titre. If antibody titres are measured close in time, such that they are in fact stable, then conditioning on starting with the maximum titre will lead to falls being erroneously observed. However, if the antibody titres are measured sufficiently far apart with respect to the rate of change with time this effect will become less important. To quantify this, we performed a sensitivity analysis in which we performed the same analysis, but only considered individuals with at least two antibody results after their maximum result and excluded the initial maximum result. This sensitivity analysis also addresses a second potential bias, where if an individual’s maximum antibody titre was measured while their levels were still climbing this may lead to the slope of the decline being under-estimated.

We used Bayesian parametric model for interval censored regression (R package, icenReg version 2.0.15) to estimate the proportion of individuals remaining antibody positive at varying times following their maximum test result allowing for the fact that antibody levels are only measured intermittently. The true event time, i.e. the time becoming antibody negative is unobserved, instead, the response interval that the event occurred within was used for the modelling.

By analysing individuals with a positive PCR test, we additionally modelled the antibody trajectory from a first positive PCR test. We used Bayesian linear mixed models as above but allowed for non-linear effects of time by using natural cubic splines, choosing the number of knots (up to 5) and subsequently their positions based on the LOOIC.

Analyses were performed using R 3.6.3 and the rstanarm library version 2.21.1. For all analyses weakly informative priors were used (see Supplementary Table S1). At least 4 chains were run per analysis to identify the burn-in period and ensure convergence, which was confirmed visually and by ensuring the Gelman-Rubin statistic was <1.1 (actual values are presented). The chains were run for a sufficient length of time to ensure the effective sample size for all parameters exceeded 200. Credibility intervals were calculated using highest posterior density intervals.

#### Role of the funding source

The funders had no role in study design; in the collection, analysis, and interpretation of data; in the writing of the report; or in the decision to submit the paper for publication.

### Supplementary tables

| Model term | Specified priors | Adjusted priors* |
| --- | --- | --- |
| Intercept | normal (1.4, 2.5) | normal (1.4, 2.9) |
| Coefficient for slope | normal (0,2.5) | normal (0, 0.05) |
| Coefficient for change in intercept (Male) |  | normal (0, 6.6) |
| Coefficient for change in slope (Male) |  | normal (0, 0.08) |
| Coefficient for change in intercept (Age) |  | normal (0, 2.4) |
| Coefficient for change in slope (Age) |  | normal (0, 0.01) |
| Coefficient for change in intercept (Black) |  | normal (0, 12.3) |
| Coefficient for change in slope (Black) |  | normal (0, 0.15) |
| Coefficient for change in intercept (Asian) |  | normal (0, 7.2) |
| Coefficient for change in slope (Asian) |  | normal (0, 0.08) |
| Coefficient for change in intercept (Other) |  | normal (0, 10.5) |
| Coefficient for change in slope (Other) |  | normal (0, 0.12) |
| Coefficient for change in intercept (Prior symptom) |  | normal (0, 5.8) |
| Coefficient for change in slope (Prior symptom) |  | normal (0, 0.05) |
| Coefficient for change in intercept (PCR-symptomatic) |  | normal (0, 7.0) |
| Coefficient for change in slope (PCR-symptomatic) |  | normal (0, 0.08) |
| Coefficient for change in intercept (PCR-asymptomatic) |  | normal (0, 8.5) |
| Coefficient for change in slope (PCR-asymptomatic) |  | normal (0, 0.10) |
| Auxiliary (sigma) | exponential (rate=1) | exponential (rate=0.88) |
| Covariance | decov (regularization = 1, concentration = 1, shape = 1, scale = 1) | decov (regularization = 1, concentration = 1, shape = 1, scale = 1) |

**Supplementary Table 1. Priors used in analysis.** *rstanarm internally adjusts the scales of the priors to make them weakly informative by default.

|  |  | Posterior mean | Monte-Carlo Standard Error (MCSE) | Posterior standard deviation (SD) | 95% Credible interval (CrI) | | Effective sample size | Gelman-Rubin statistic (Rhat) |
| --- | --- | --- | --- | --- | --- | --- | --- | --- |
| Baseline model | Intercept | 2.08917 | 0.00085 | 0.03241 | 2.02481 | 2.15160 | 1455 | 1.00 |
|  | t (slope) | -0.01172 | 0.00001 | 0.00031 | -0.01232 | -0.01111 | 2262 | 1.00 |
|  | Standard deviation of errors (sigma) | 0.28047 | 0.00064 | 0.01317 | 0.25588 | 0.30860 | 427 | 1.01 |
|  | Random effect intercept standard deviation | 0.39783 | 0.00088 | 0.03089 | 0.34109 | 0.46177 | 1241 | 1.00 |
|  | Random effect intercept & slope covariance | 0.00258 | 0.00000 | 0.00024 | 0.00214 | 0.00306 | 2236 | 1.00 |
|  | Random effect slope standard deviation | 0.00003 | 0.00000 | 0.00000 | 0.00002 | 0.00004 | 952 | 1.00 |
| Gender model | Intercept: Female | 2.08429 | 0.00113 | 0.03637 | 2.01404 | 2.15667 | 1031 | 1.00 |
|  | t (slope): Female | -0.01179 | 0.00001 | 0.00035 | -0.01248 | -0.01108 | 2010 | 1.00 |
|  | Change in intercept: Male | 0.00830 | 0.00227 | 0.07497 | -0.13754 | 0.15534 | 1087 | 1.00 |
|  | Change in slope: Male | 0.00024 | 0.00002 | 0.00073 | -0.00116 | 0.00166 | 1844 | 1.00 |
|  | Standard deviation of errors (sigma) | 0.28035 | 0.00080 | 0.01460 | 0.25264 | 0.30968 | 336 | 1.01 |
|  | Random effect intercept standard deviation | 0.39923 | 0.00094 | 0.03213 | 0.33895 | 0.46301 | 1176 | 1.00 |
|  | Random effect intercept & slope covariance | 0.00258 | 0.00001 | 0.00025 | 0.00212 | 0.00309 | 1984 | 1.00 |
|  | Random effect slope standard deviation | 0.00003 | 0.00000 | 0.00000 | 0.00002 | 0.00004 | 696 | 1.00 |
| Age model | Intercept :41-years old | 1.93419 | 0.00289 | 0.11256 | 1.83195 | 2.03702 | 1517 | 1.00 |
|  | t (slope): 41-years old | -0.01331 | 0.00002 | 0.00107 | -0.01434 | -0.01228 | 2439 | 1.00 |
|  | Change in intercept: per 10-year older | 0.09246 | 0.00069 | 0.02697 | 0.03743 | 0.14368 | 1525 | 1.00 |
|  | Change in slope: per 10-year older | 0.00095 | 0.00001 | 0.00025 | 0.00045 | 0.00144 | 2466 | 1.00 |
|  | Standard deviation of errors (sigma) | 0.27913 | 0.00080 | 0.01430 | 0.25278 | 0.30950 | 319 | 1.01 |
|  | Random effect intercept standard deviation | 0.38754 | 0.00095 | 0.03137 | 0.33221 | 0.45420 | 1092 | 1.00 |
|  | Random effect intercept & slope covariance | 0.00246 | 0.00001 | 0.00023 | 0.00201 | 0.00294 | 1808 | 1.00 |
|  | Random effect slope standard deviation | 0.00003 | 0.00000 | 0.00000 | 0.00002 | 0.00003 | 719 | 1.00 |
| Ethnicity model | Intercept: White | 2.00937 | 0.00116 | 0.03871 | 1.93435 | 2.08513 | 1109 | 1.00 |
|  | t (slope): White | -0.01229 | 0.00001 | 0.00038 | -0.01304 | -0.01154 | 1884 | 1.00 |
|  | Change in intercept: Black | 0.15572 | 0.00320 | 0.13793 | -0.11145 | 0.42489 | 1854 | 1.00 |
|  | Change in intercept: Asian | 0.26813 | 0.00227 | 0.08109 | 0.10807 | 0.42587 | 1274 | 1.00 |
|  | Change in intercept: Other | 0.19450 | 0.00311 | 0.11963 | -0.03466 | 0.42948 | 1476 | 1.00 |
|  | Change in slope: Black | 0.00083 | 0.00003 | 0.00137 | -0.00193 | 0.00349 | 2767 | 1.00 |
|  | Change in slope: Asian | 0.00183 | 0.00002 | 0.00081 | 0.00021 | 0.00340 | 2376 | 1.00 |
|  | Change in slope: Other | 0.00179 | 0.00002 | 0.00116 | -0.00047 | 0.00409 | 2583 | 1.00 |
|  | Standard deviation of errors (sigma) | 0.28016 | 0.00072 | 0.01385 | 0.25438 | 0.30888 | 371 | 1.02 |
|  | Random effect intercept standard deviation | 0.38866 | 0.00083 | 0.03110 | 0.33174 | 0.45090 | 1398 | 1.00 |
|  | Random effect intercept & slope covariance | 0.00250 | 0.00000 | 0.00024 | 0.00204 | 0.00298 | 2469 | 1.00 |
|  | Random effect slope standard deviation | 0.00003 | 0.00000 | 0.00000 | 0.00002 | 0.00004 | 824 | 1.01 |
| Prior symptom model | Intercept: No | 1.96200 | 0.00133 | 0.04976 | 1.86363 | 2.05783 | 1405 | 1.00 |
|  | t (slope): No | -0.01199 | 0.00001 | 0.00049 | -0.01292 | -0.01104 | 2210 | 1.00 |
|  | Change in intercept: Yes | 0.20553 | 0.00164 | 0.06471 | 0.08020 | 0.33126 | 1558 | 1.00 |
|  | Change in slope: Yes | 0.00040 | 0.00001 | 0.00064 | -0.00081 | 0.00162 | 2237 | 1.00 |
|  | Standard deviation of errors (sigma) | 0.27967 | 0.00100 | 0.01474 | 0.25312 | 0.31050 | 217 | 1.03 |
|  | Random effect intercept standard deviation | 0.38960 | 0.00104 | 0.03174 | 0.33133 | 0.45671 | 923 | 1.01 |
|  | Random effect intercept & slope covariance | 0.00256 | 0.00001 | 0.00024 | 0.00211 | 0.00303 | 2180 | 1.00 |
|  | Random effect slope standard deviation | 0.00003 | 0.00000 | 0.00000 | 0.00002 | 0.00004 | 492 | 1.01 |
| PCR model | Intercept: No | 2.03221 | 0.00103 | 0.03955 | 1.95242 | 2.11065 | 1473 | 1.00 |
|  | t (slope): No | -0.01248 | 0.00001 | 0.00039 | -0.01325 | -0.01173 | 2362 | 1.00 |
|  | Change in intercept: Symptomatic | 0.21074 | 0.00211 | 0.07966 | 0.05619 | 0.36638 | 1420 | 1.00 |
|  | Change in intercept: Asymptomatic | 0.07506 | 0.00243 | 0.09633 | -0.11404 | 0.26730 | 1567 | 1.00 |
|  | Change in slope: Symptomatic | 0.00241 | 0.00002 | 0.00078 | 0.00097 | 0.00400 | 2207 | 1.00 |
|  | Change in slope: Asymptomatic | 0.00174 | 0.00002 | 0.00092 | -0.00005 | 0.00355 | 2674 | 1.00 |
|  | Standard deviation of errors (sigma) | 0.27887 | 0.00079 | 0.01420 | 0.25269 | 0.30766 | 324 | 1.01 |
|  | Random effect intercept standard deviation | 0.39308 | 0.00089 | 0.03153 | 0.33412 | 0.45938 | 1263 | 1.00 |
|  | Random effect intercept & slope covariance | 0.00250 | 0.00001 | 0.00024 | 0.00205 | 0.00299 | 2095 | 1.00 |
|  | Random effect slope standard deviation | 0.00003 | 0.00000 | 0.00000 | 0.00002 | 0.00004 | 742 | 1.00 |
| Multivariable model | Intercept | 1.73447 | 0.00304 | 0.11633 | 1.59589 | 1.87703 | 1465 | 1.00 |
|  | t (slope) | -0.01433 | 0.00002 | 0.00115 | -0.01569 | -0.01295 | 2353 | 1.00 |
|  | Change in intercept: Male | -0.02490 | 0.00187 | 0.07344 | -0.17412 | 0.11775 | 1547 | 1.00 |
|  | Change in intercept: per 10-year older | 0.08209 | 0.00073 | 0.02647 | 0.02996 | 0.13387 | 1314 | 1.00 |
|  | Change in intercept: Black | 0.16392 | 0.00334 | 0.13832 | -0.11071 | 0.43263 | 1715 | 1.00 |
|  | Change in intercept: Asian | 0.24471 | 0.00229 | 0.08123 | 0.08251 | 0.40655 | 1259 | 1.00 |
|  | Change in intercept: Other | 0.19820 | 0.00280 | 0.11583 | -0.03129 | 0.42335 | 1712 | 1.00 |
|  | Change in intercept: Had symptom | 0.18587 | 0.00162 | 0.06487 | 0.05610 | 0.30973 | 1598 | 1.00 |
|  | Change in intercept: Symptomatic PCR | 0.13758 | 0.00230 | 0.08069 | -0.01969 | 0.29519 | 1233 | 1.00 |
|  | Change in intercept: Asymptomatic PCR | 0.06518 | 0.00263 | 0.09549 | -0.12199 | 0.25202 | 1316 | 1.00 |
|  | Change in slope: Male | -0.00014 | 0.00001 | 0.00073 | -0.00158 | 0.00128 | 2435 | 1.00 |
|  | Change in slope: per 10-year older | 0.00089 | 0.00001 | 0.00026 | 0.00036 | 0.00139 | 2266 | 1.00 |
|  | Change in slope: Black | 0.00056 | 0.00003 | 0.00135 | -0.00204 | 0.00321 | 2501 | 1.00 |
|  | Change in slope: Asian | 0.00149 | 0.00002 | 0.00081 | -0.00011 | 0.00305 | 2575 | 1.00 |
|  | Change in slope: Other | 0.00187 | 0.00002 | 0.00113 | -0.00028 | 0.00407 | 2572 | 1.00 |
|  | Change in slope: Had symptom | 0.00007 | 0.00001 | 0.00064 | -0.00119 | 0.00135 | 2719 | 1.00 |
|  | Change in slope: Symptomatic PCR | 0.00217 | 0.00002 | 0.00078 | 0.00059 | 0.00369 | 2551 | 1.00 |
|  | Change in slope: Asymptomatic PCR | 0.00151 | 0.00002 | 0.00089 | -0.00023 | 0.00329 | 2184 | 1.00 |
|  | Standard deviation of errors (sigma) | 0.27831 | 0.00070 | 0.01404 | 0.25289 | 0.30824 | 398 | 1.01 |
|  | Random effect intercept standard deviation | 0.37052 | 0.00080 | 0.03067 | 0.31432 | 0.43385 | 1462 | 1.00 |
|  | Random effect intercept & slope covariance | 0.00233 | 0.00000 | 0.00023 | 0.00191 | 0.00278 | 2186 | 1.00 |
|  | Random effect slope standard deviation | 0.00003 | 0.00000 | 0.00000 | 0.00002 | 0.00003 | 765 | 1.00 |

**Supplementary Table S2. Original model coefficients and MCMC diagnostics for univariable and multivariable models.** In the multivariable model the following characteristics are used as baseline: age 41 years, gender female, ethnicity white, no prior symptoms and no positive PCR test.

| **Indication** | **Ct value** | **Platform** | **Maximum Abbott titre** |
| --- | --- | --- | --- |
| Asymptomatic | 24.8 | Abbott RealTime | 0.01 |
| Asymptomatic | 24.8 | Abbott RealTime | 0.01 |
| Asymptomatic | 31.2 | Abbott RealTime | 0.01 |
| Asymptomatic | 30.6 | Altona RealStar | 0.01 |
| Asymptomatic | 30.1 | Abbott RealTime | 0.02 |
| Asymptomatic | 26.6 | Abbott RealTime | 0.02 |
| Asymptomatic | 26.3 | Abbott RealTime | 0.02 |
| Asymptomatic | 31.2 | Abbott RealTime | 0.02 |
| Asymptomatic | 31.1 | Abbott RealTime | 0.02 |
| Asymptomatic | 30.1 | Abbott RealTime | 0.02 |
| Asymptomatic | 26.0 | Abbott RealTime | 0.02 |
| Asymptomatic | 30.4 | Abbott RealTime | 0.03 |
| Asymptomatic | 37.0 | ABI 7500 RealTime | 0.03 |
| Asymptomatic | 30.0 | Abbott RealTime | 0.04 |
| Asymptomatic | 29.7 | Abbott RealTime | 0.04 |
| Asymptomatic | 31.3 | Abbott RealTime | 0.04 |
| Asymptomatic | 36.3 | ABI 7500 RealTime | 0.04 |
| Asymptomatic | 28.5 | Abbott RealTime | 0.05 |
| Asymptomatic | 26.6 | Abbott RealTime | 0.06 |
| Asymptomatic | 29.6 | Abbott RealTime | 0.07 |
| Asymptomatic | 24.2 | Abbott RealTime | 0.07 |
| Asymptomatic | 10.3 | Abbott RealTime | 0.07 |
| Asymptomatic | 30.2 | Abbott RealTime | 0.07 |
| Asymptomatic | 14.1 | Altona RealStar | 0.24 |
| Asymptomatic | 26.8 | Abbott RealTime | 0.25 |
| Asymptomatic | 37.3 | ABI 7500 RealTime | 0.3 |
| Asymptomatic | 8.8 | Abbott RealTime | 0.38 |
| Asymptomatic | 20.8 | Abbott RealTime | 0.61 |
| Asymptomatic | 21.4 | Abbott RealTime | 0.61 |
| Asymptomatic | 20.6 | Abbott RealTime | 1.04 |
| Asymptomatic | 24.5 | Abbott RealTime | 1.33 |
| Asymptomatic | 30.6 | Abbott RealTime | 1.36 |
| Asymptomatic | 28.9 | Abbott RealTime | 1.36 |
| Asymptomatic | 30.9 | Altona RealStar | 1.45 |
| Asymptomatic | 19.7 | Abbott RealTime | 1.49 |
| Asymptomatic | 27.2 | Altona RealStar | 1.64 |
| Asymptomatic | 24.6 | Abbott RealTime | 1.7 |
| Asymptomatic | 25.3 | Abbott RealTime | 1.8 |
| Asymptomatic | 20.7 | Abbott RealTime | 1.87 |
| Asymptomatic | 13.5 | Abbott RealTime | 1.95 |
| Asymptomatic | 4.0 | Abbott RealTime | 2.08 |
| Asymptomatic | 6.9 | Abbott RealTime | 2.1 |
| Asymptomatic | 27.3 | Abbott RealTime | 2.13 |
| Asymptomatic | 28.8 | Abbott RealTime | 2.15 |
| Asymptomatic | 4.9 | Abbott RealTime | 2.18 |
| Asymptomatic | 29.6 | Abbott RealTime | 2.3 |
| Asymptomatic | 26.8 | Altona RealStar | 2.32 |
| Asymptomatic | 27.4 | Abbott RealTime | 2.62 |
| Asymptomatic | 26.6 | Abbott RealTime | 2.85 |
| Asymptomatic | 22.2 | Abbott RealTime | 3.03 |
| Asymptomatic | 7.3 | Abbott RealTime | 3.17 |
| Asymptomatic | 18.8 | Abbott RealTime | 3.24 |
| Asymptomatic | 26.5 | Abbott RealTime | 3.39 |
| Asymptomatic | 27.2 | Abbott RealTime | 3.45 |
| Asymptomatic | 28.1 | Abbott RealTime | 3.49 |
| Asymptomatic | 28.5 | Abbott RealTime | 3.51 |
| Asymptomatic | 23.3 | Abbott RealTime | 3.58 |
| Asymptomatic | 6.2 | Abbott RealTime | 3.73 |
| Asymptomatic | 25.2 | Abbott RealTime | 3.82 |
| Asymptomatic | 17.4 | Abbott RealTime | 3.83 |
| Asymptomatic | 26.3 | Abbott RealTime | 3.95 |
| Asymptomatic | 26.4 | Abbott RealTime | 4.11 |
| Asymptomatic | 4.1 | Abbott RealTime | 4.16 |
| Asymptomatic | 26.4 | Abbott RealTime | 4.16 |
| Asymptomatic | 26.8 | Abbott RealTime | 4.23 |
| Asymptomatic | 19.2 | Abbott RealTime | 4.33 |
| Asymptomatic | 9.0 | Abbott RealTime | 4.34 |
| Asymptomatic | 11.2 | Abbott RealTime | 4.39 |
| Asymptomatic | 30.3 | Abbott RealTime | 4.48 |
| Asymptomatic | 23.8 | Abbott RealTime | 4.59 |
| Asymptomatic | 29.7 | Abbott RealTime | 4.6 |
| Asymptomatic | 23.2 | Abbott RealTime | 4.78 |
| Asymptomatic | 25.1 | Abbott RealTime | 4.83 |
| Asymptomatic | 5.5 | Abbott RealTime | 4.86 |
| Asymptomatic | 20.9 | Abbott RealTime | 4.87 |
| Asymptomatic | 24.9 | Abbott RealTime | 4.89 |
| Asymptomatic | 24.0 | Abbott RealTime | 5 |
| Asymptomatic | 22.9 | Abbott RealTime | 5.09 |
| Asymptomatic | 16.6 | Abbott RealTime | 5.15 |
| Asymptomatic | 21.1 | Abbott RealTime | 5.21 |
| Asymptomatic | 22.0 | Abbott RealTime | 5.24 |
| Asymptomatic | 24.3 | Abbott RealTime | 5.33 |
| Asymptomatic | 28.0 | Abbott RealTime | 5.57 |
| Asymptomatic | 22.1 | Abbott RealTime | 5.62 |
| Asymptomatic | 21.7 | Abbott RealTime | 5.63 |
| Asymptomatic | 28.2 | Abbott RealTime | 5.78 |
| Asymptomatic | 21.9 | Abbott RealTime | 5.83 |
| Asymptomatic | 26.3 | Abbott RealTime | 5.92 |
| Asymptomatic | 21.9 | Abbott RealTime | 5.95 |
| Asymptomatic | 22.7 | Abbott RealTime | 6.15 |
| Asymptomatic | 23.9 | Abbott RealTime | 6.37 |
| Asymptomatic | 24.1 | Abbott RealTime | 6.37 |
| Asymptomatic | 20.4 | Abbott RealTime | 6.65 |
| Asymptomatic | 21.8 | Abbott RealTime | 6.78 |
| Asymptomatic | 18.7 | Altona RealStar | 6.82 |
| Asymptomatic | 19.0 | Abbott RealTime | 6.85 |
| Asymptomatic | 25.2 | Abbott RealTime | 6.89 |
| Asymptomatic | 15.0 | Abbott RealTime | 7.03 |
| Asymptomatic | 26.9 | Abbott RealTime | 7.04 |
| Asymptomatic | 26.8 | Abbott RealTime | 7.05 |
| Asymptomatic | 26.1 | Abbott RealTime | 7.19 |
| Asymptomatic | 25.4 | Abbott RealTime | 7.26 |
| Asymptomatic | 22.7 | Altona RealStar | 7.34 |
| Asymptomatic | 27.1 | Abbott RealTime | 7.51 |
| Asymptomatic | 23.0 | Abbott RealTime | 7.55 |
| Asymptomatic | 20.1 | Abbott RealTime | 7.63 |
| Asymptomatic | 29.8 | Abbott RealTime | 7.82 |
| Asymptomatic | 25.2 | Abbott RealTime | 7.83 |
| Symptomatic | 31.8 | Altona RealStar | 0.01 |
| Symptomatic | 28.1 | Abbott RealTime | 0.04 |
| Symptomatic | 16.6 | Altona RealStar | 0.04 |
| Symptomatic | 18.4 | Abbott RealTime | 0.07 |
| Symptomatic | 26.3 | Altona RealStar | 0.09 |
| Symptomatic | 9.6 | Abbott RealTime | 0.19 |
| Symptomatic | 22.9 | RdRp | 0.39 |
| Symptomatic | 11.3 | Altona RealStar | 0.97 |
| Symptomatic | 30.5 | RdRp | 1.12 |
| Symptomatic | 23.3 | RdRp | 1.14 |
| Symptomatic | 5.0 | Abbott RealTime | 1.15 |
| Symptomatic | 30.3 | RdRp | 1.29 |
| Symptomatic | 19.5 | RdRp | 1.43 |
| Symptomatic | 29.4 | RdRp | 1.58 |
| Symptomatic | 18.0 | RdRp | 1.69 |
| Symptomatic | 9.8 | Abbott RealTime | 1.75 |
| Symptomatic | 19.1 | RdRp | 1.85 |
| Symptomatic | 5.2 | Abbott RealTime | 1.86 |
| Symptomatic | 19.7 | RdRp | 1.89 |
| Symptomatic | 18.6 | Abbott RealTime | 2.09 |
| Symptomatic | 20.7 | RdRp | 2.21 |
| Symptomatic | 20.1 | Abbott RealTime | 2.36 |
| Symptomatic | 21.6 | Abbott RealTime | 2.41 |
| Symptomatic | 20.4 | RdRp | 2.49 |
| Symptomatic | 32.0 | RdRp | 2.57 |
| Symptomatic | 20.6 | RdRp | 2.63 |
| Symptomatic | 15.7 | Altona RealStar | 2.76 |
| Symptomatic | 21.5 | Altona RealStar | 2.76 |
| Symptomatic | 17.1 | RdRp | 2.84 |
| Symptomatic | 20.7 | RdRp | 2.9 |
| Symptomatic | 18.3 | RdRp | 3.2 |
| Symptomatic | 3.5 | Abbott RealTime | 3.28 |
| Symptomatic | 21.5 | RdRp | 3.34 |
| Symptomatic | 24.2 | RdRp | 3.52 |
| Symptomatic | 9.4 | Abbott RealTime | 3.63 |
| Symptomatic | 17.0 | RdRp | 3.72 |
| Symptomatic | 22.4 | RdRp | 3.75 |
| Symptomatic | 19.0 | RdRp | 3.79 |
| Symptomatic | 20.3 | RdRp | 3.79 |
| Symptomatic | 6.6 | Abbott RealTime | 3.85 |
| Symptomatic | 29.9 | RdRp | 3.91 |
| Symptomatic | 11.8 | Abbott RealTime | 3.92 |
| Symptomatic | 16.7 | Altona RealStar | 3.93 |
| Symptomatic | 25.1 | RdRp | 4.05 |
| Symptomatic | 24.4 | RdRp | 4.24 |
| Symptomatic | 30.3 | RdRp | 4.29 |
| Symptomatic | 17.5 | Altona RealStar | 4.36 |
| Symptomatic | 32.2 | RdRp | 4.39 |
| Symptomatic | 19.7 | RdRp | 4.48 |
| Symptomatic | 19.6 | RdRp | 4.51 |
| Symptomatic | 7.8 | Abbott RealTime | 4.52 |
| Symptomatic | 20.2 | RdRp | 4.57 |
| Symptomatic | 22.1 | RdRp | 4.77 |
| Symptomatic | 35.2 | RdRp | 4.85 |
| Symptomatic | 21.8 | RdRp | 4.9 |
| Symptomatic | 23.5 | Altona RealStar | 4.91 |
| Symptomatic | 25.1 | RdRp | 4.92 |
| Symptomatic | 18.5 | Altona RealStar | 4.99 |
| Symptomatic | 22.7 | RdRp | 5.04 |
| Symptomatic | 27.4 | RdRp | 5.07 |
| Symptomatic | 23.1 | Altona RealStar | 5.22 |
| Symptomatic | 28.8 | RdRp | 5.24 |
| Symptomatic | 18.7 | RdRp | 5.24 |
| Symptomatic | 16.8 | RdRp | 5.3 |
| Symptomatic | 21.8 | Abbott RealTime | 5.32 |
| Symptomatic | 29.7 | RdRp | 5.33 |
| Symptomatic | 25.5 | RdRp | 5.35 |
| Symptomatic | 31.3 | RdRp | 5.4 |
| Symptomatic | 22.8 | RdRp | 5.43 |
| Symptomatic | 21.1 | RdRp | 5.46 |
| Symptomatic | 26.7 | RdRp | 5.65 |
| Symptomatic | 19.7 | RdRp | 5.65 |
| Symptomatic | 26.3 | RdRp | 5.68 |
| Symptomatic | 25.2 | Abbott RealTime | 5.71 |
| Symptomatic | 12.2 | Abbott RealTime | 5.76 |
| Symptomatic | 26.4 | RdRp | 5.77 |
| Symptomatic | 22.5 | RdRp | 5.77 |
| Symptomatic | 20.9 | RdRp | 5.94 |
| Symptomatic | 18.1 | RdRp | 6 |
| Symptomatic | 19.8 | RdRp | 6.04 |
| Symptomatic | 25.1 | RdRp | 6.07 |
| Symptomatic | 27.8 | RdRp | 6.13 |
| Symptomatic | 25.7 | RdRp | 6.19 |
| Symptomatic | 26.6 | RdRp | 6.25 |
| Symptomatic | 21.5 | RdRp | 6.25 |
| Symptomatic | 23.2 | RdRp | 6.31 |
| Symptomatic | 17.9 | RdRp | 6.31 |
| Symptomatic | 22.0 | RdRp | 6.37 |
| Symptomatic | 18.8 | RdRp | 6.38 |
| Symptomatic | 16.0 | Abbott RealTime | 6.38 |
| Symptomatic | 24.3 | RdRp | 6.4 |
| Symptomatic | 21.4 | RdRp | 6.48 |
| Symptomatic | 21.4 | RdRp | 6.54 |
| Symptomatic | 12.3 | Abbott RealTime | 6.55 |
| Symptomatic | 16.7 | Abbott RealTime | 6.64 |
| Symptomatic | 19.6 | RdRp | 6.66 |
| Symptomatic | 21.6 | Abbott RealTime | 6.69 |
| Symptomatic | 18.1 | RdRp | 6.72 |
| Symptomatic | 19.4 | RdRp | 6.73 |
| Symptomatic | 18.9 | RdRp | 6.78 |
| Symptomatic | 26.9 | RdRp | 6.79 |
| Symptomatic | 26.1 | Abbott RealTime | 6.79 |
| Symptomatic | 17.5 | RdRp | 6.81 |
| Symptomatic | 23.5 | Altona RealStar | 6.82 |
| Symptomatic | 25.4 | RdRp | 6.83 |
| Symptomatic | 13.0 | Abbott RealTime | 6.91 |
| Symptomatic | 27.9 | RdRp | 6.97 |
| Symptomatic | 20.3 | RdRp | 7 |
| Symptomatic | 27.7 | RdRp | 7.04 |
| Symptomatic | 30.1 | RdRp | 7.06 |
| Symptomatic | 12.0 | Altona RealStar | 7.08 |
| Symptomatic | 16.7 | RdRp | 7.24 |
| Symptomatic | 19.3 | RdRp | 7.31 |
| Symptomatic | 30.6 | RdRp | 7.36 |
| Symptomatic | 23.2 | RdRp | 7.39 |
| Symptomatic | 36.0 | RdRp | 7.52 |
| Symptomatic | 20.3 | RdRp | 7.52 |
| Symptomatic | 31.5 | RdRp | 7.58 |
| Symptomatic | 36.3 | RdRp | 7.67 |
| Symptomatic | 28.4 | RdRp | 7.72 |
| Symptomatic | 16.8 | RdRp | 7.78 |
| Symptomatic | 23.9 | Abbott RealTime | 7.85 |
| Symptomatic | 19.1 | Abbott RealTime | 8.08 |
| Symptomatic | 19.0 | RdRp | 8.39 |

**Supplementary Table S3. PCR cycle threshold (Ct) values and corresponding maximum Abbott CMIA IgG antibody titres.** The Ct values obtained across different platforms are not directly comparable. The median Ct values for the two most commonly used PCR platforms Abbott (n=123) and Public Health England’s RdRp assay (n=86) were higher in those who did not seroconvert (maximum antibody titre<1.40) vs. those that did, 26.7 vs. 22.1 (Kruskal-Wallis p=0.0002) and 26.8 vs. 21.9 (p=0.10).

### Supplementary File legend

Observed serial IgG titres are plotted for 452 healthcare workers, timed from the first available antibody result. Antibody results occurring before the maximum titre are shown in blue and were excluded from the fitted model. The fitted model is overlaid, with the posterior mean trajectory indicated by the solid line and the 3 intensities of shaded areas indicating the 50%, 80% and 95% credible intervals.

### Supplementary Figures

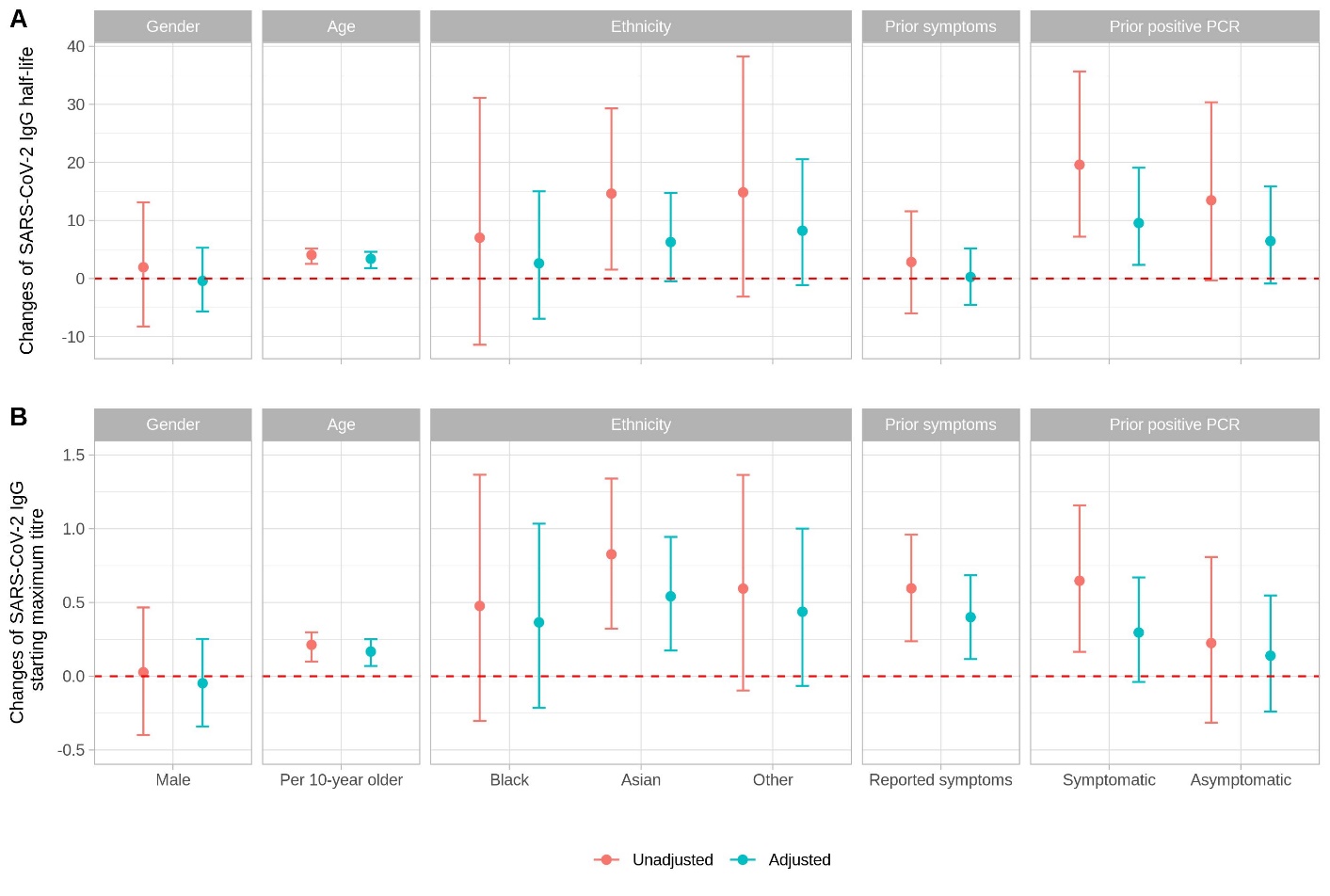

**Figure S1. Comparison of the changes of SARS-CoV-2 IgG half-life (Panel A) and starting maximum titre (Panel B) in the univariable (unadjusted) and multivariable (adjusted) models**. The baseline group for gender is female, for ethnicity is white, for symptoms is no reported prior symptoms and for prior PCR results is no prior positive. The dashed horizontal line indicates no effect.

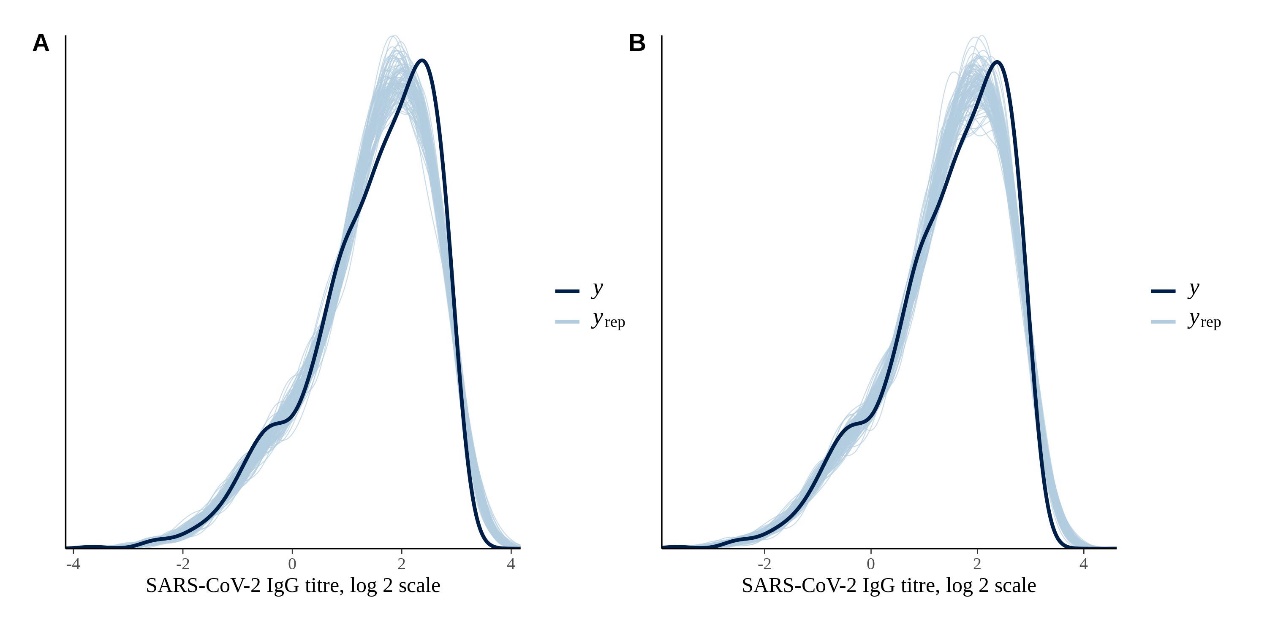

**Figure S2. Posterior predictive check for the baseline model (Panel A) and the multivariable model (Panel B).** The plot compares the distribution of the observed SARS-CoV-2 IgG titres on a log2 scale (y) and 100 simulated datasets drawn from the posterior predictive distribution (yrep). From the plot, the model is able to generate data that resembles the observed data.

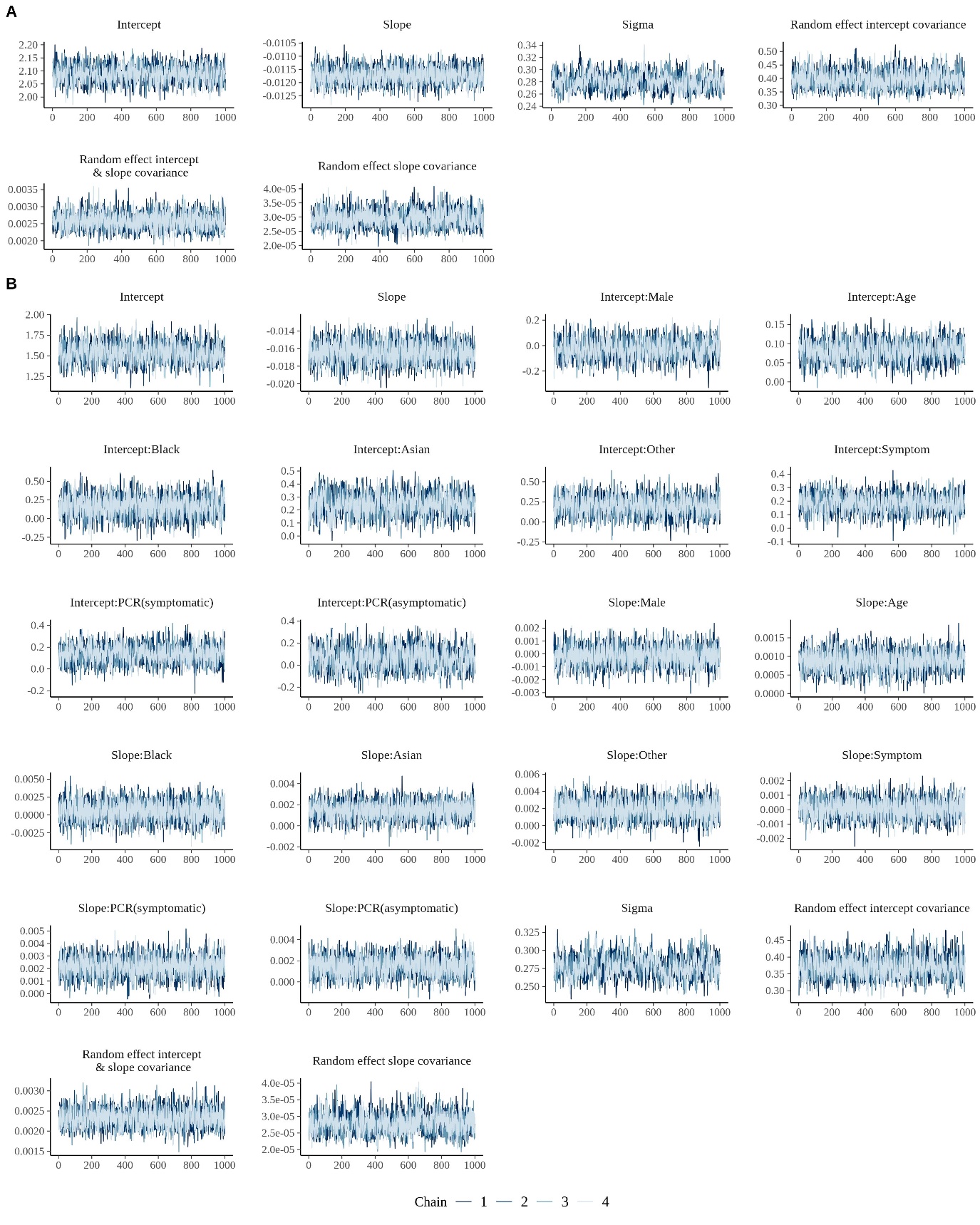

**Figure S3. MCMC trace plots for assessing convergence of chains.** Panel A shows the trace plots for the baseline model. Panel B shows the trace plots for the multivariable model. In both sets of plots a burn in period of 1000 iterations has been discarded prior to plotting.

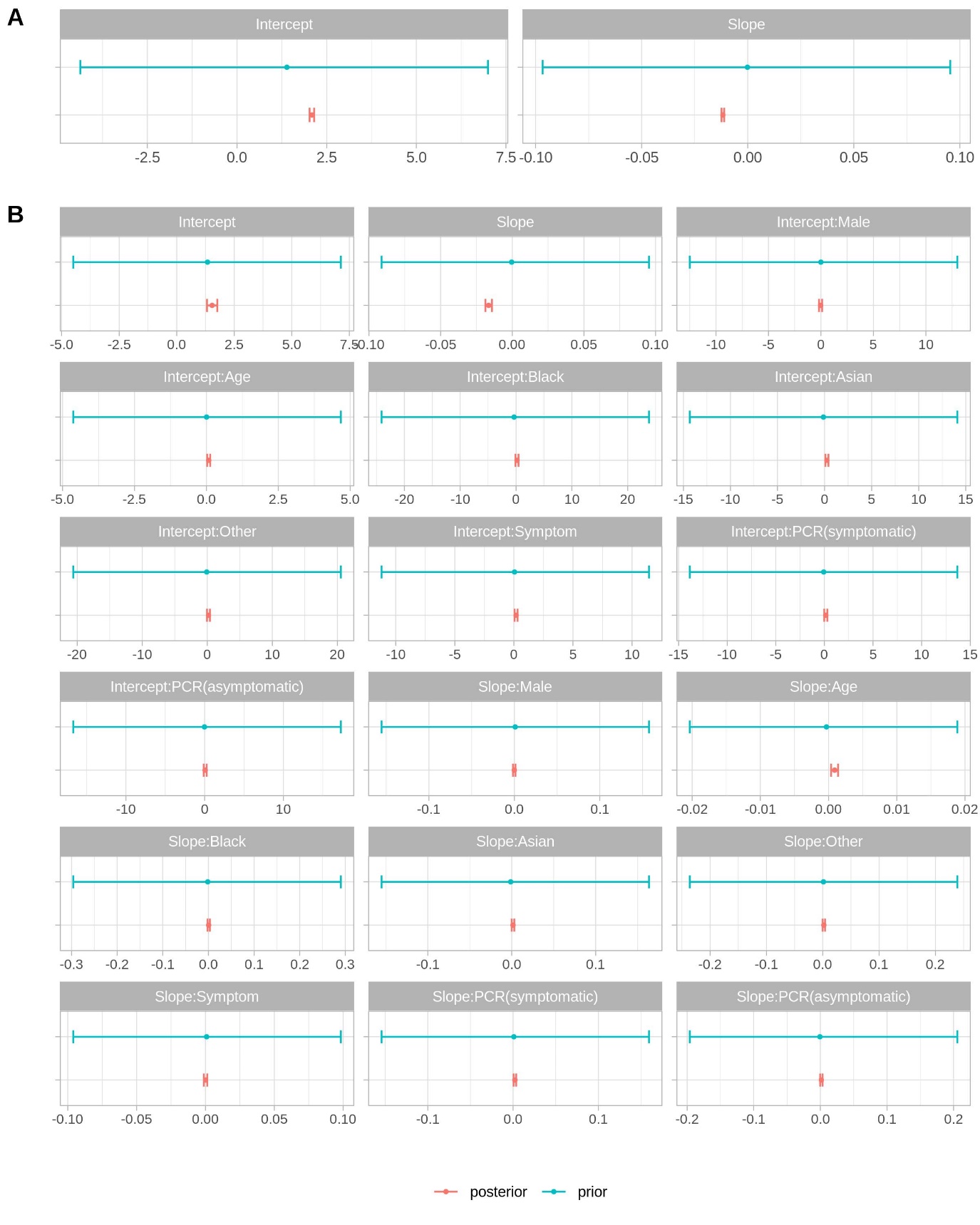

**Figure S4. Comparison of the mean and 95% highest density intervals of model parameter prior and posterior distributions.** Panel A shows the intercept and slope in the baseline model. Panel B shows the intercept, slope, and covariates coefficients in the multivariable model.

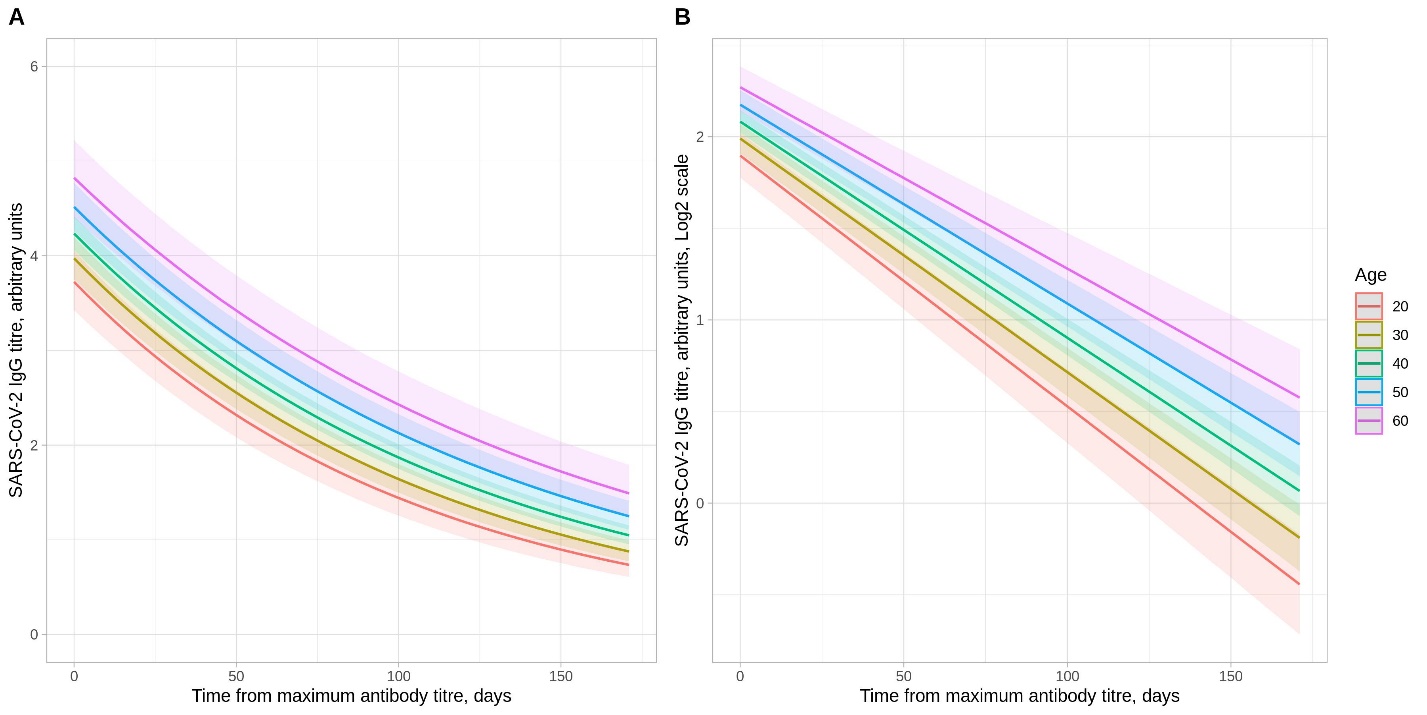

**Figure S5. Posterior mean trajectories of SARS-CoV-2 IgG after maximum IgG antibody level by age categories**. Panel A shows the relationship on an untransformed scale and Panel B using a log2 scale.

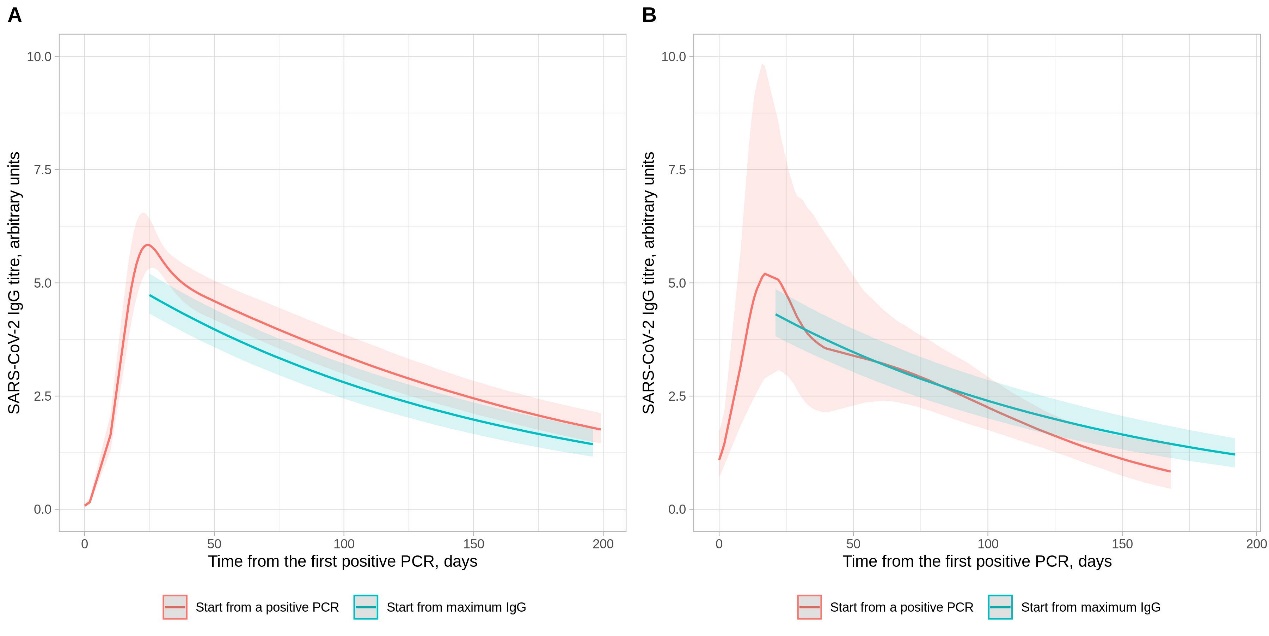

**Figure S6. Comparison of SARS-CoV-2 IgG antibody levels following a positive PCR test and the maximum IgG level per individual in those with a positive PCR test**. Panel A shows those with a positive PCR undertaken for symptoms; Panel B shows those with a positive PCR for asymptomatic screening. For plotting purposes, the x-axis value for the model starting from the maximum IgG level is aligned to the maximum point from the model starting with a positive PCR test. The model starting from a positive PCR is fitted with a 5-knot spline (3 interior knots at t=10, t=30, and t=50).
