## Supplementary File for "The duration, dynamics and determinants of SARS-CoV-2 antibody responses in individual healthcare workers"

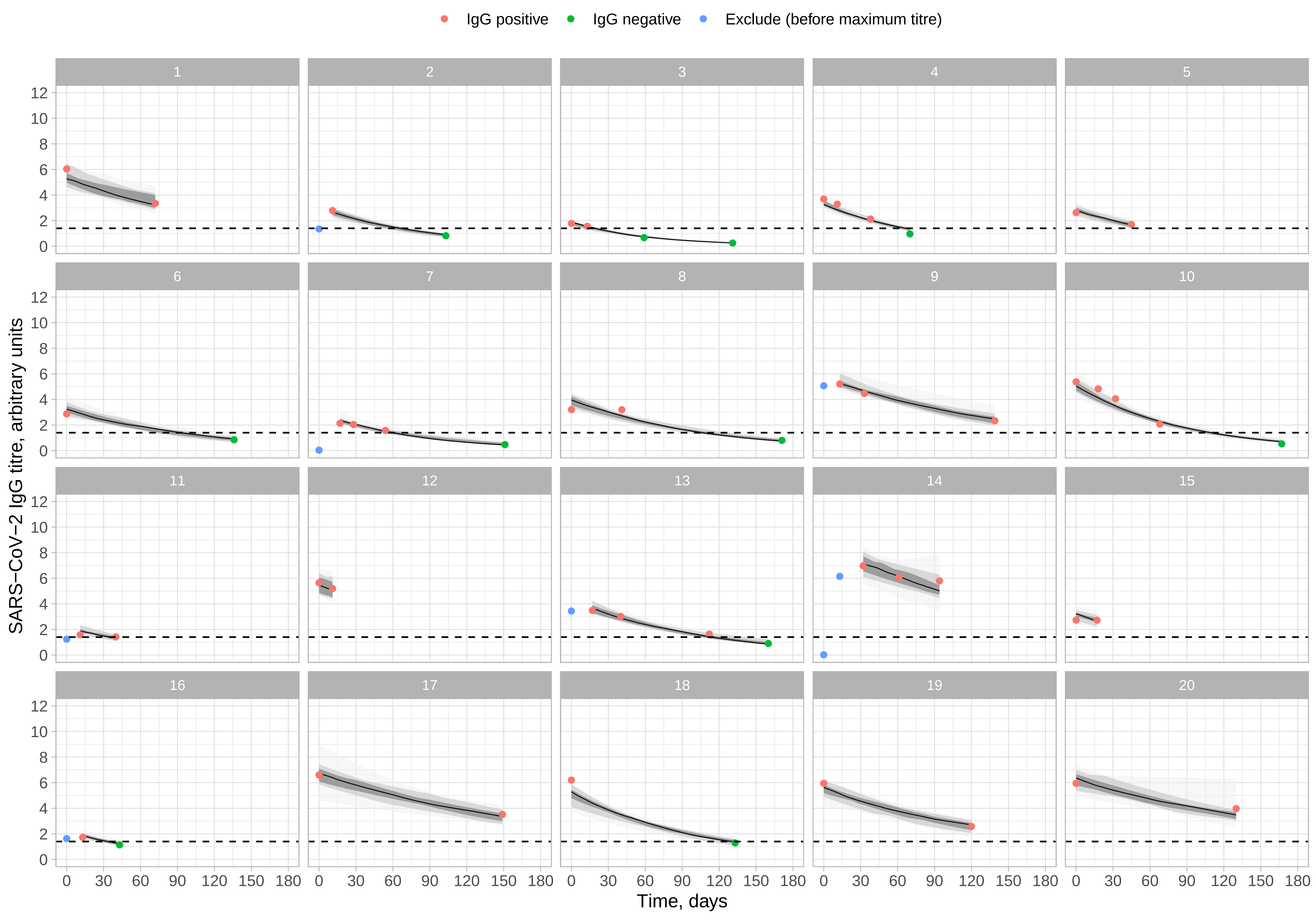

IgG positive
IgG negative
Exclude (before maximum titre)
SARS−CoV−2 IgG titre, arbitrary units
Time, days
0
30
60
90
120
150
180
0
2
4
6
8
10
12

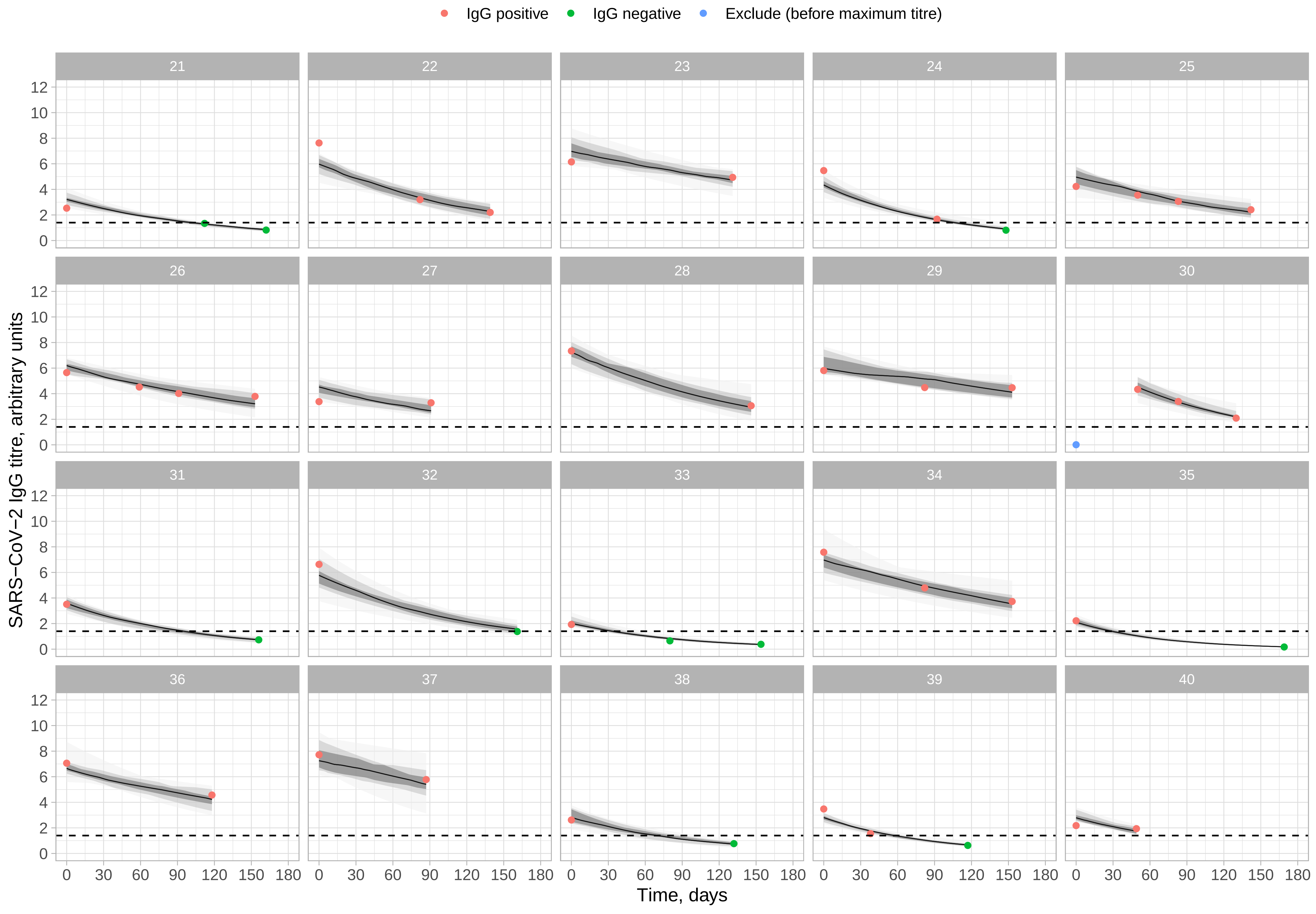

IgG positive
IgG negative
Exclude (before maximum titre)
SARS−CoV−2 IgG titre, arbitrary units
Time, days
0
2
4
6
8
10
12
0
30
60
90
120
150
180

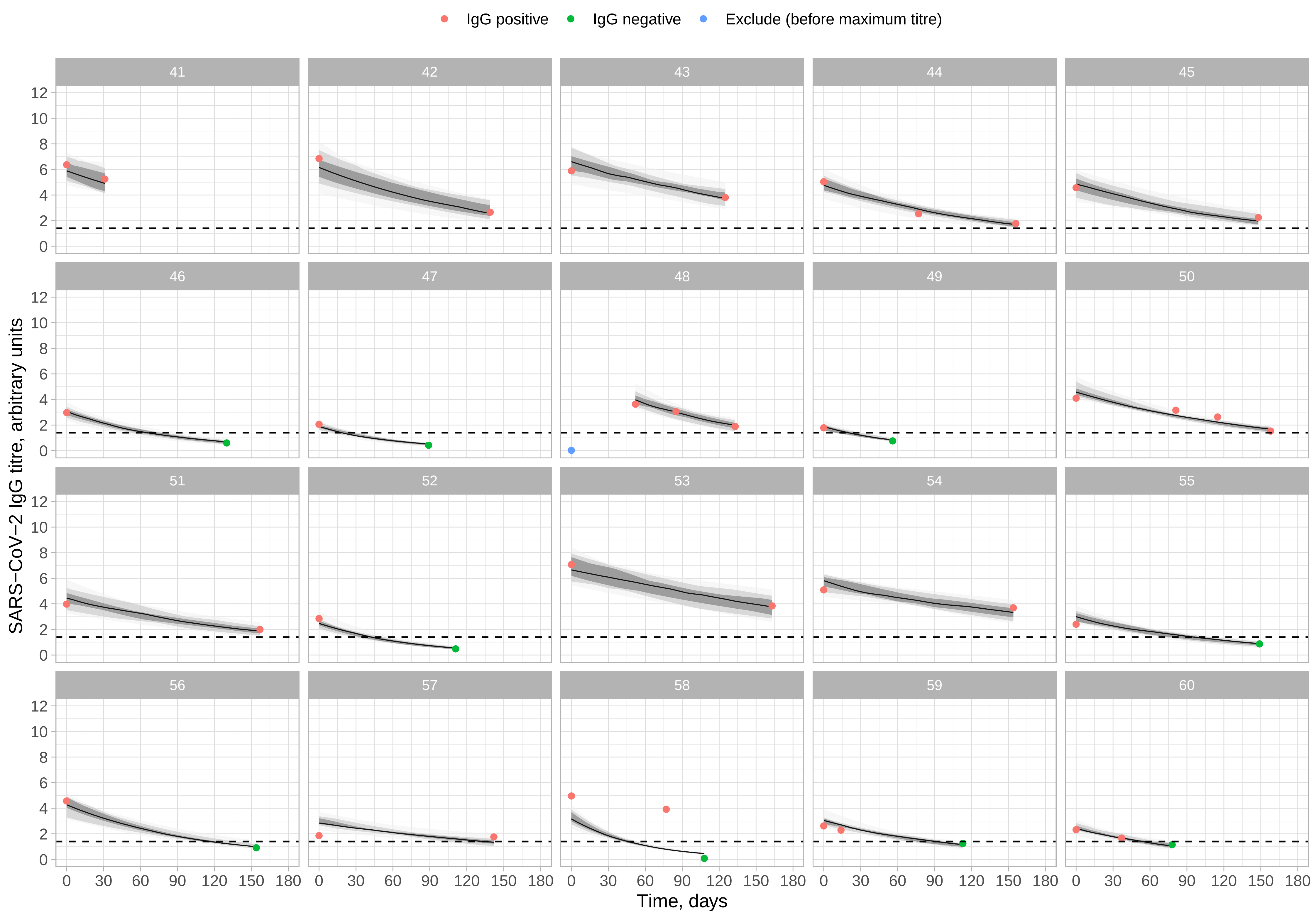

IgG positive
IgG negative
Exclude (before maximum titre)
SARS−CoV−2 IgG titre, arbitrary units
Time, days
0
2
4
6
8
10
12
0
30
60
90
120
150
180

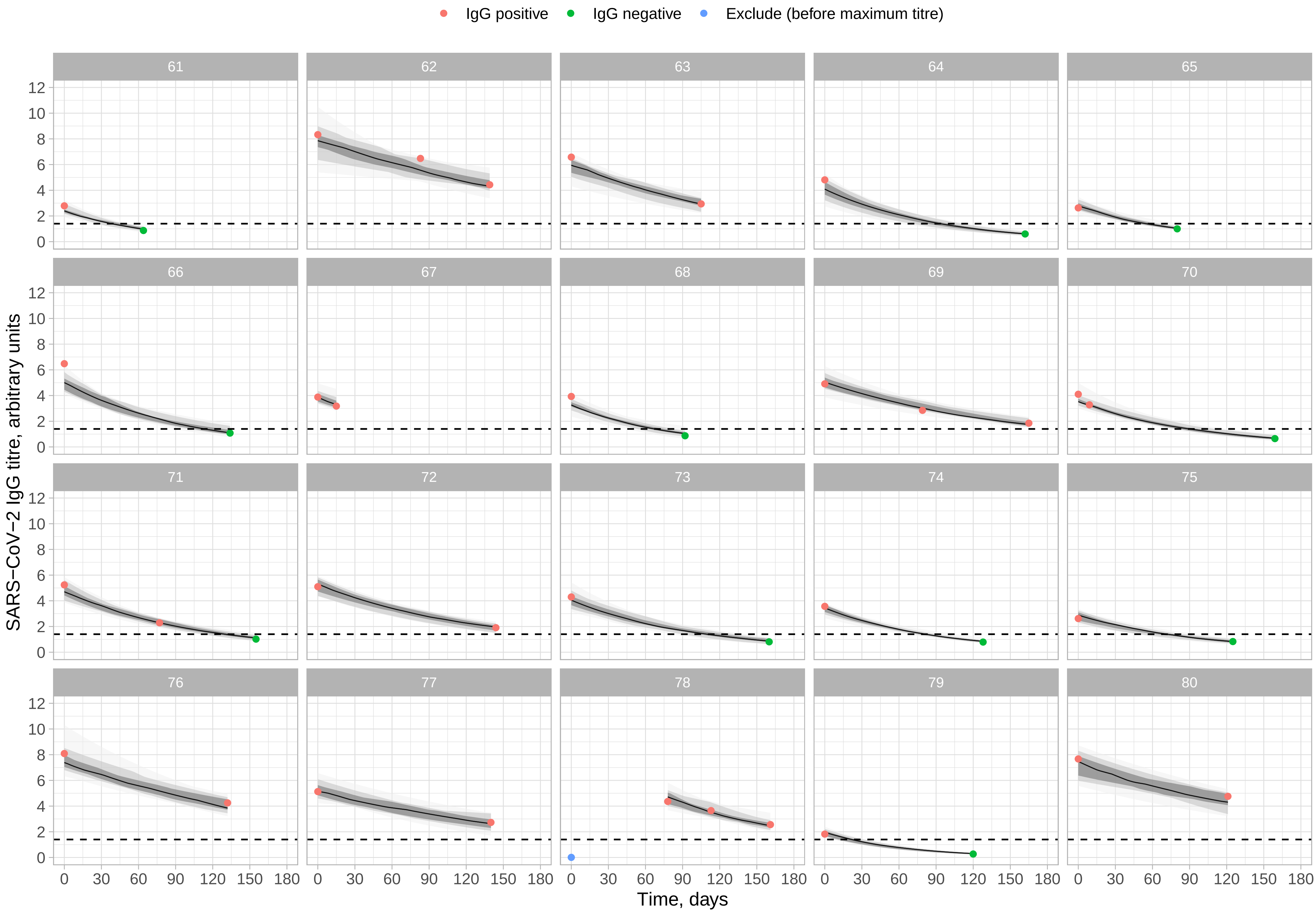

IgG positive
IgG negative
Exclude (before maximum titre)
SARS−CoV−2 IgG titre, arbitrary units
0
2
4
6
8
10
12
0
30
60
90
120
150
180
Time, days

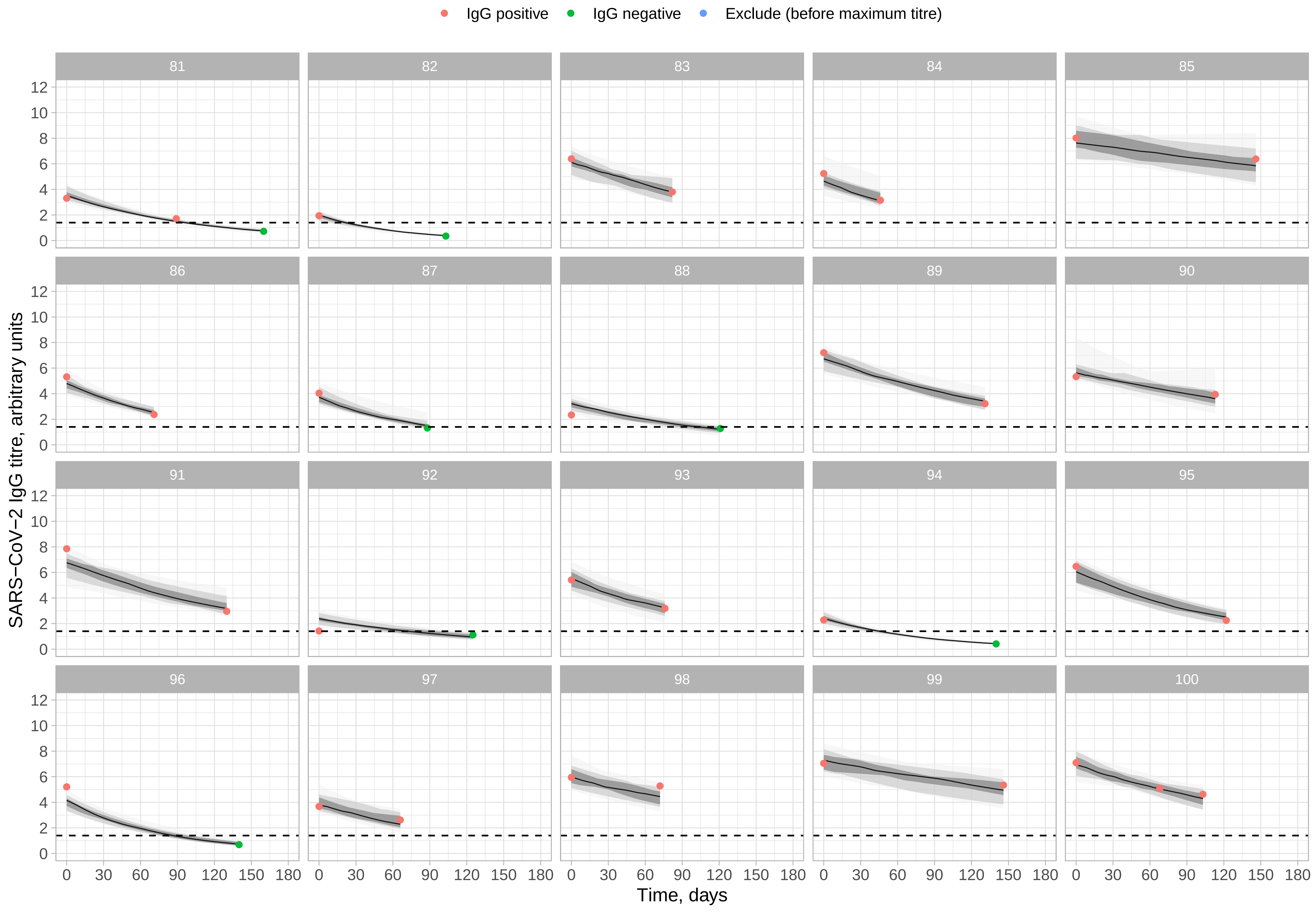

IgG positive
IgG negative
Exclude (before maximum titre)
SARS−CoV−2 IgG titre, arbitrary units
Time, days
0
2
4
6
8
10
12
0
30
60
90
120
150
180

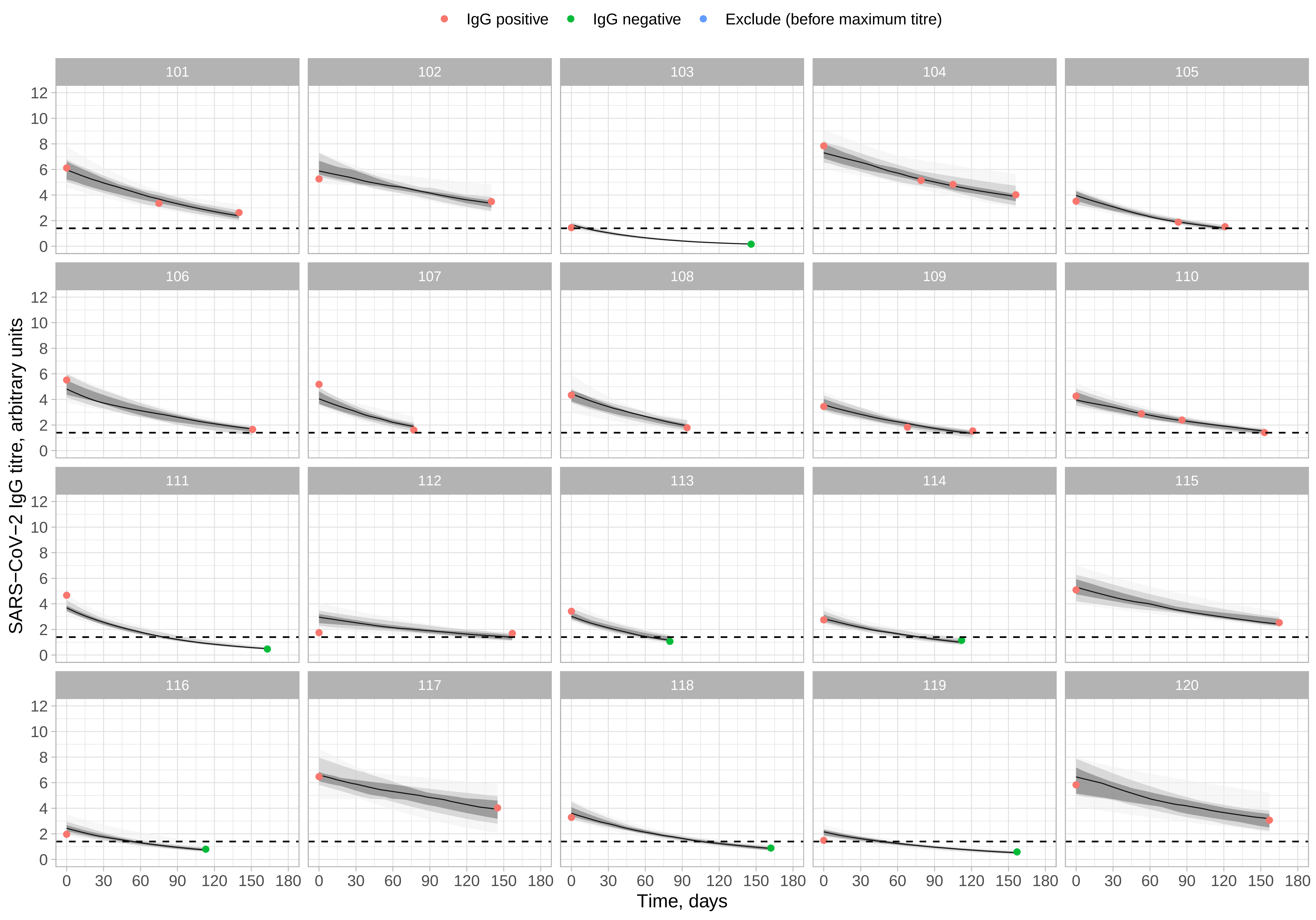
IgG positive
IgG negative
Exclude (before maximum titre)
SARS−CoV−2 IgG titre, arbitrary units
Time, days
0
2
4
6
8
10
12
0
30
60
90
120
150
180

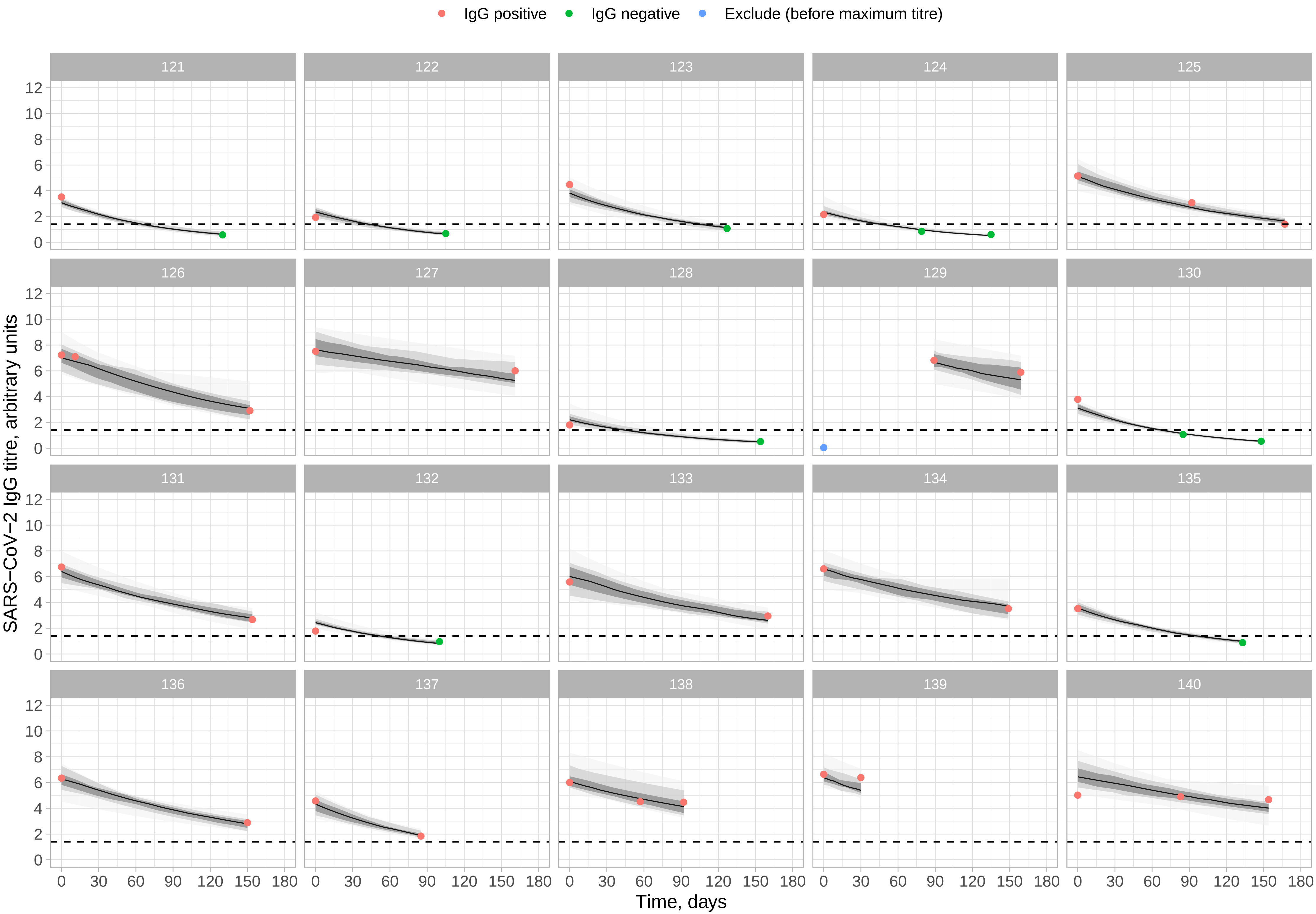

IgG positive
IgG negative
Exclude (before maximum titre)
SARS−CoV−2 IgG titre, arbitrary units
Time, days
0
30
60
90
120
150
180
0
2
4
6
8
10
12

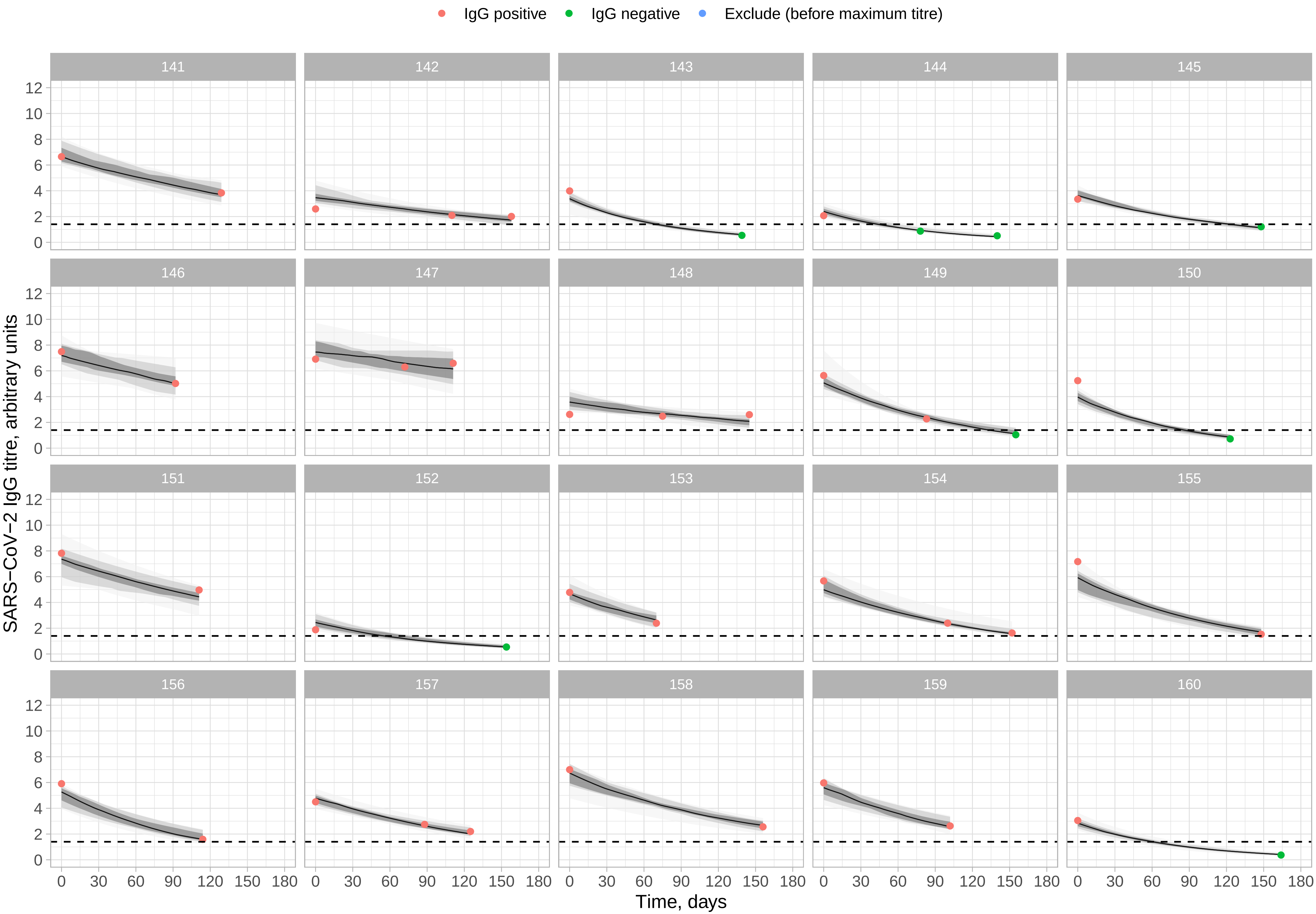
IgG positive
IgG negative
Exclude (before maximum titre)
SARS−CoV−2 IgG titre, arbitrary units
Time, days
0
2
4
6
8
10
12
0
30
60
90
120
150
180

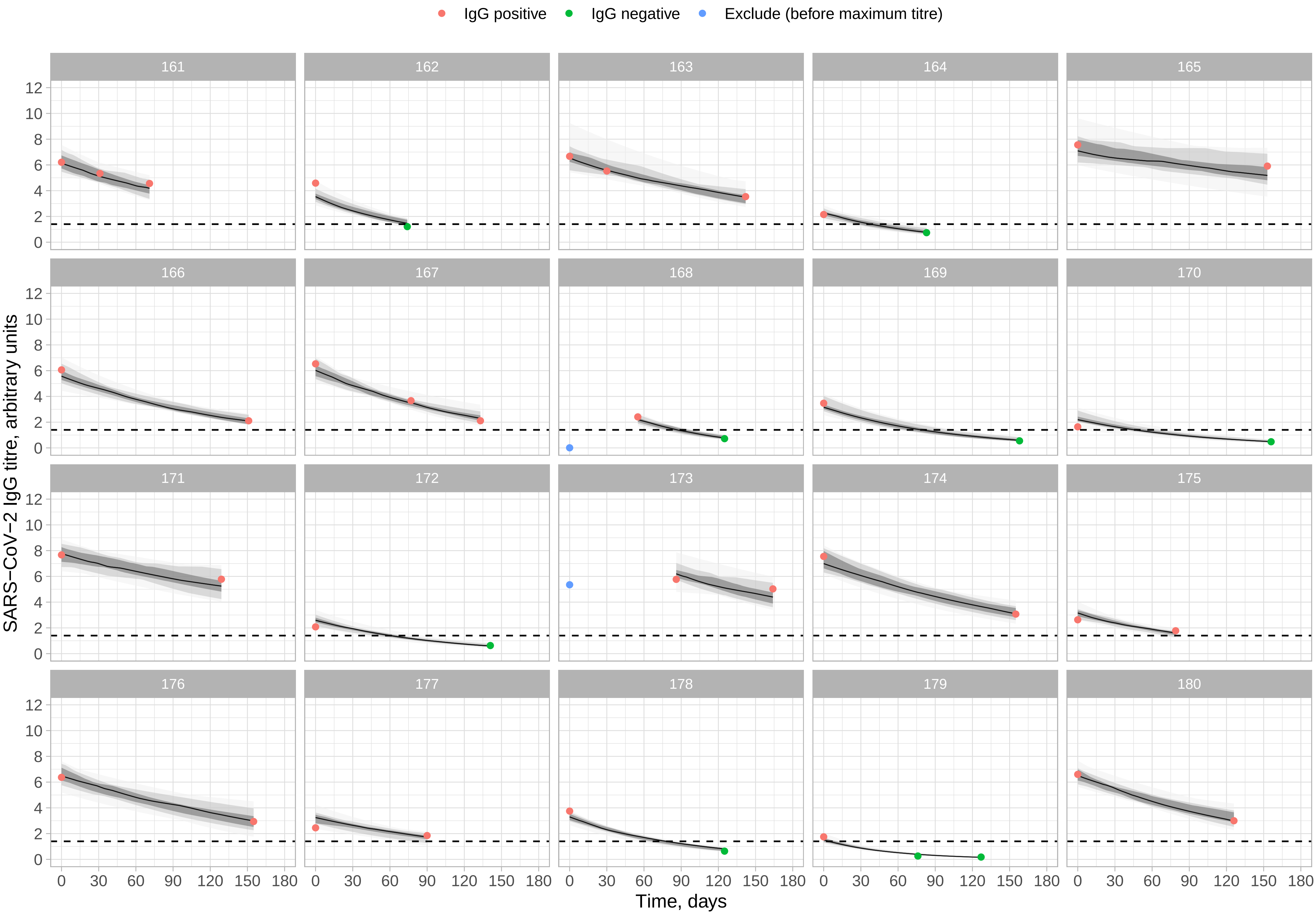

IgG positive
IgG negative
Exclude (before maximum titre)
SARS−CoV−2 IgG titre, arbitrary units
Time, days
0
2
4
6
8
10
12
0
30
60
90
120
150
180

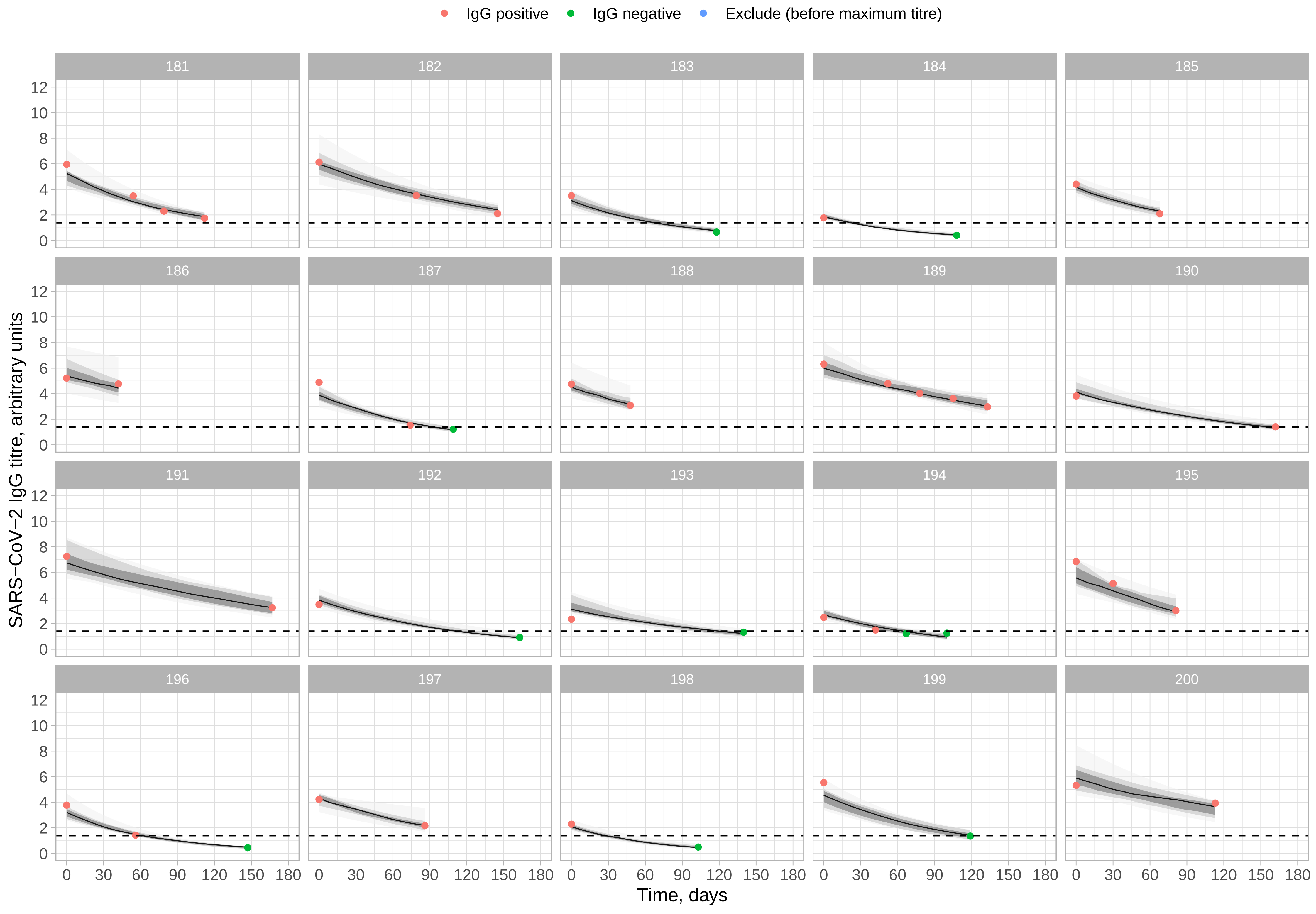
IgG positive
IgG negative
Exclude (before maximum titre)
SARS−CoV−2 IgG titre, arbitrary units
Time, days
0
2
4
6
8
10
12
0
30
60
90
120
150
180

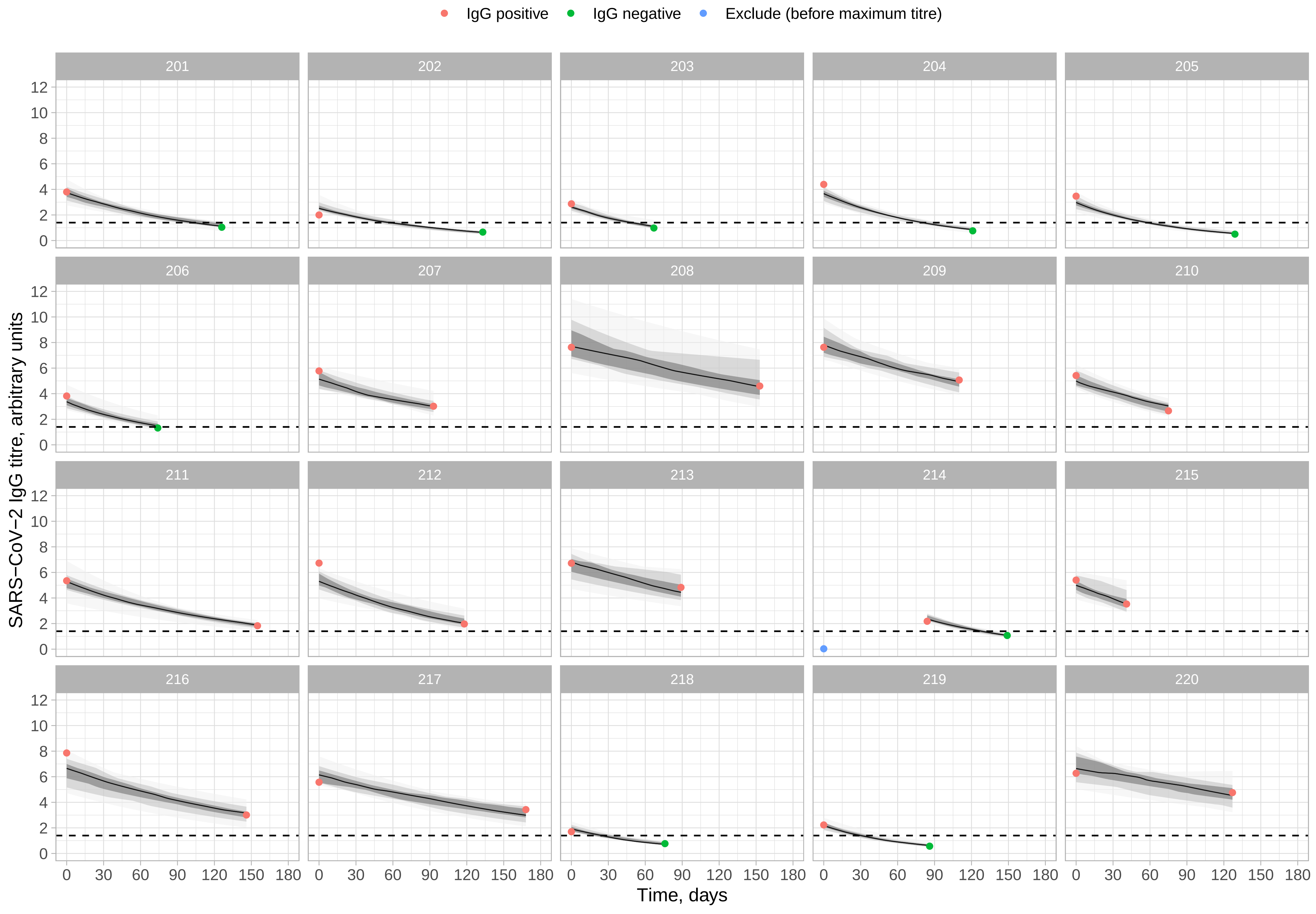

IgG positive
IgG negative
Exclude (before maximum titre)
SARS−CoV−2 IgG titre, arbitrary units
Time, days
0
2
4
6
8
10
12
0
30
60
90
120
150
180

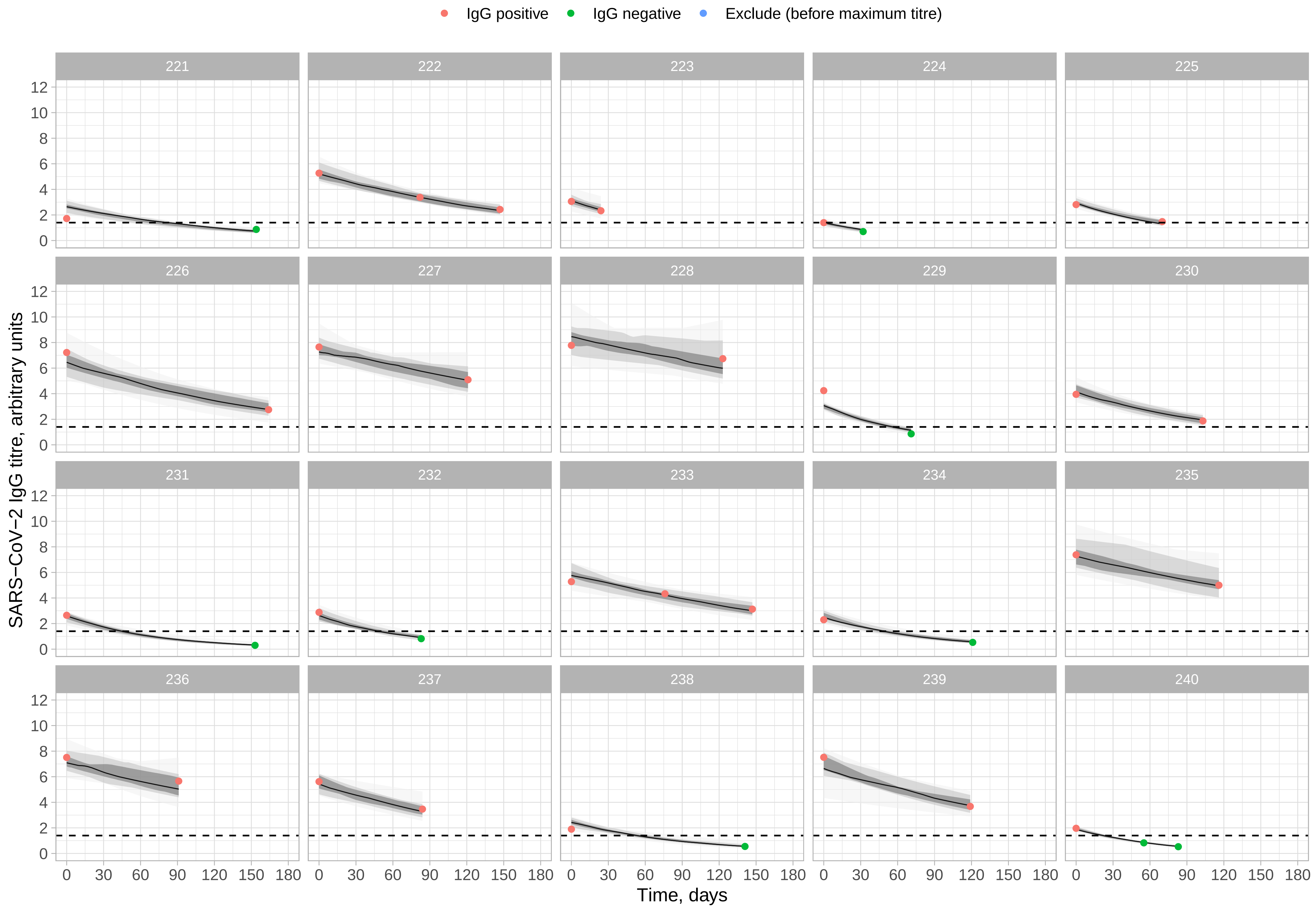

IgG positive
IgG negative
Exclude (before maximum titre)
SARS−CoV−2 IgG titre, arbitrary units
0
2
4
6
8
10
12
0
30
60
90
120
150
180
Time, days

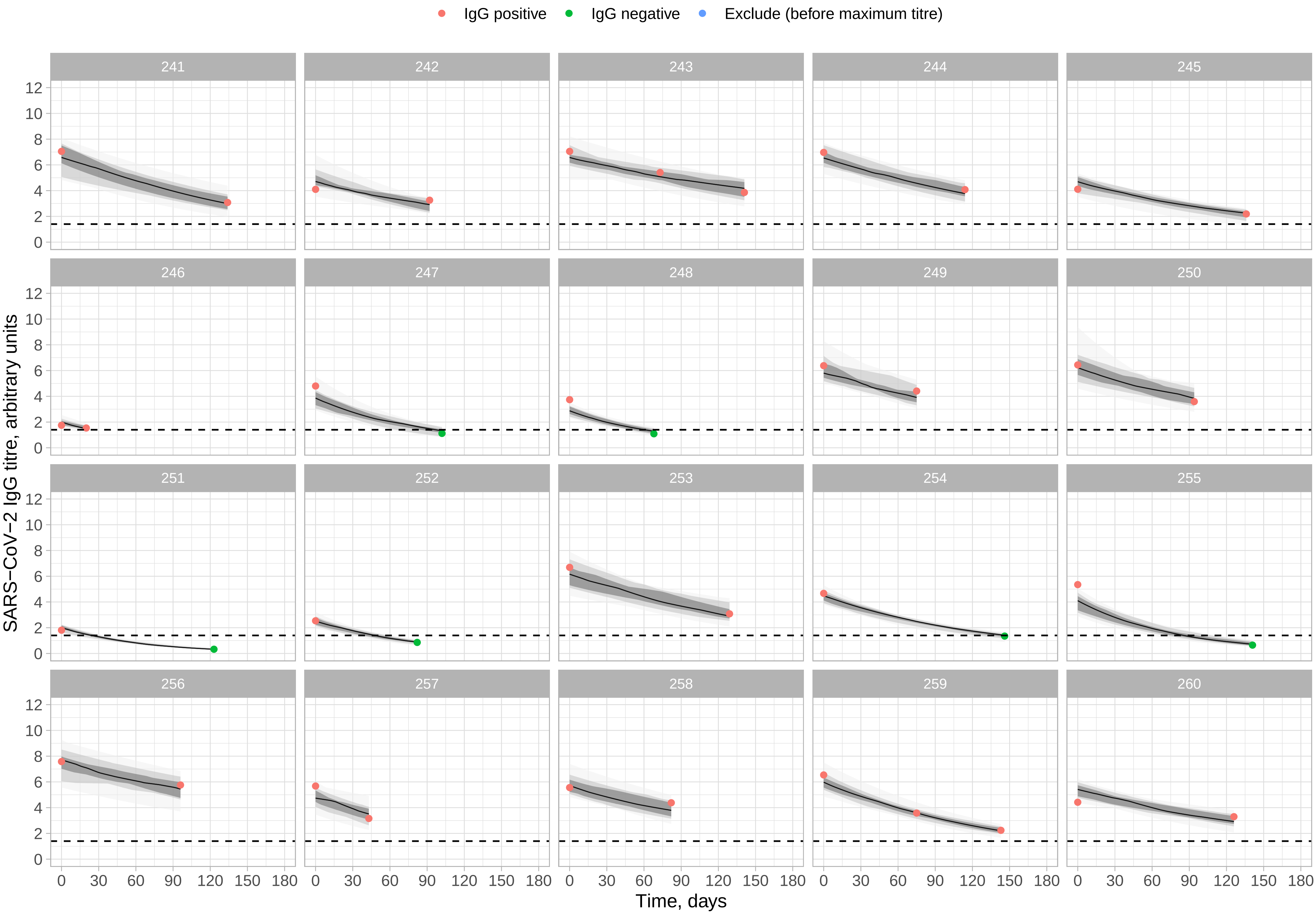
IgG positive
IgG negative
Exclude (before maximum titre)
SARS−CoV−2 IgG titre, arbitrary units
Time, days

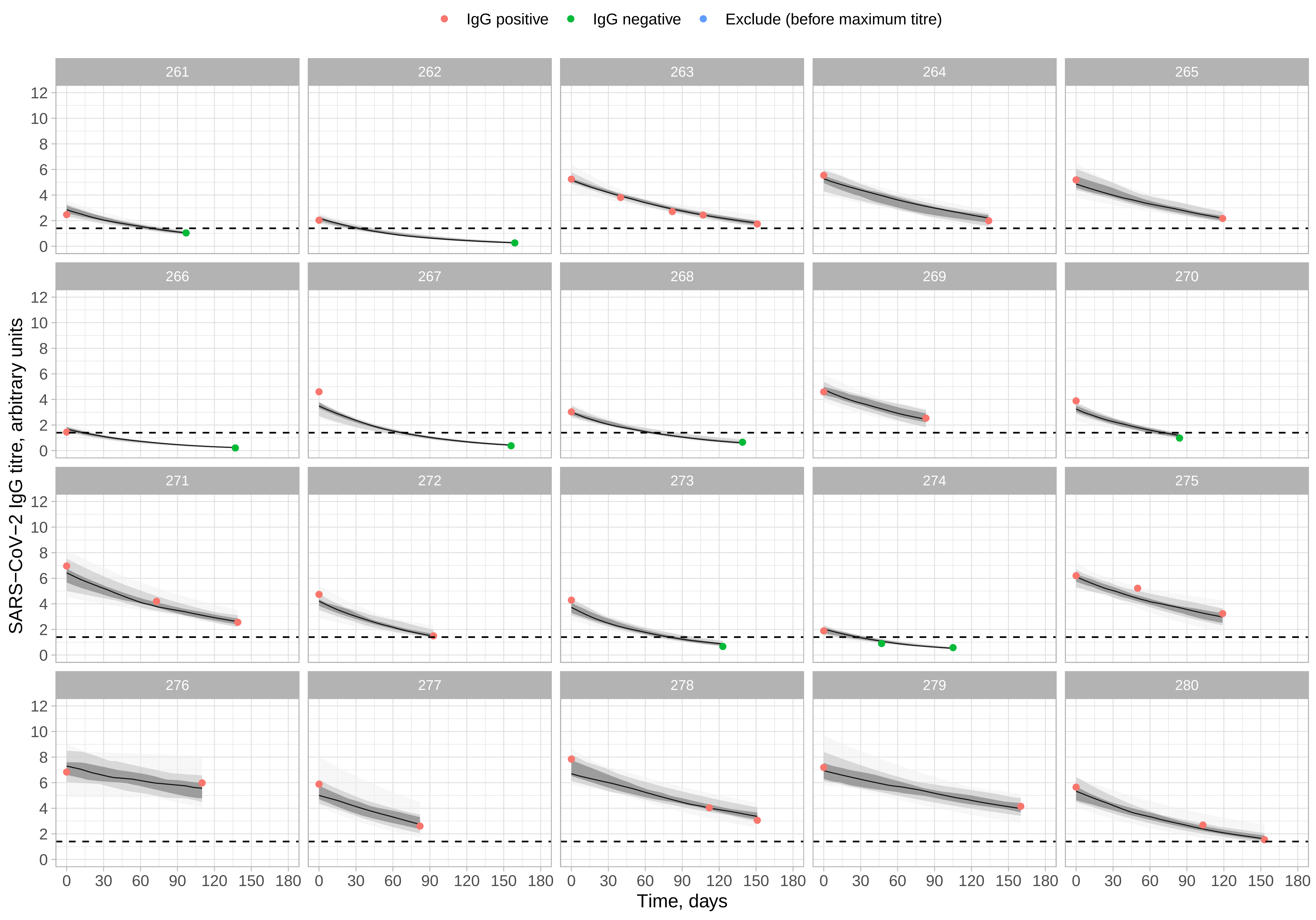

IgG positive
IgG negative
Exclude (before maximum titre)
SARS−CoV−2 IgG titre, arbitrary units
0
2
4
6
8
10
12
0
30
60
90
120
150
180
Time, days

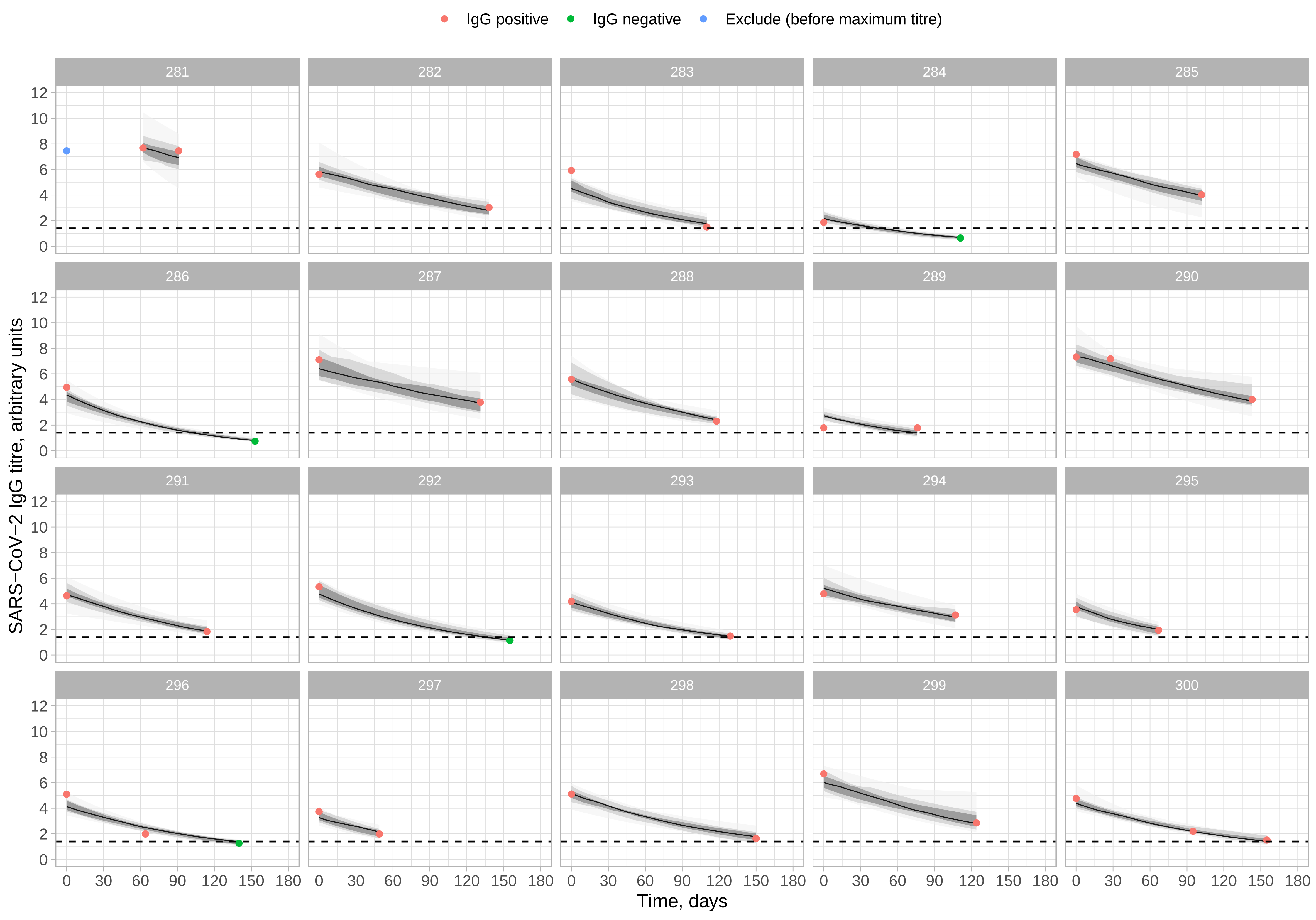

IgG positive
IgG negative
Exclude (before maximum titre)
SARS−CoV−2 IgG titre, arbitrary units
Time, days
0 2 4 6 8 10 12
0 30 60 90 120 150 180

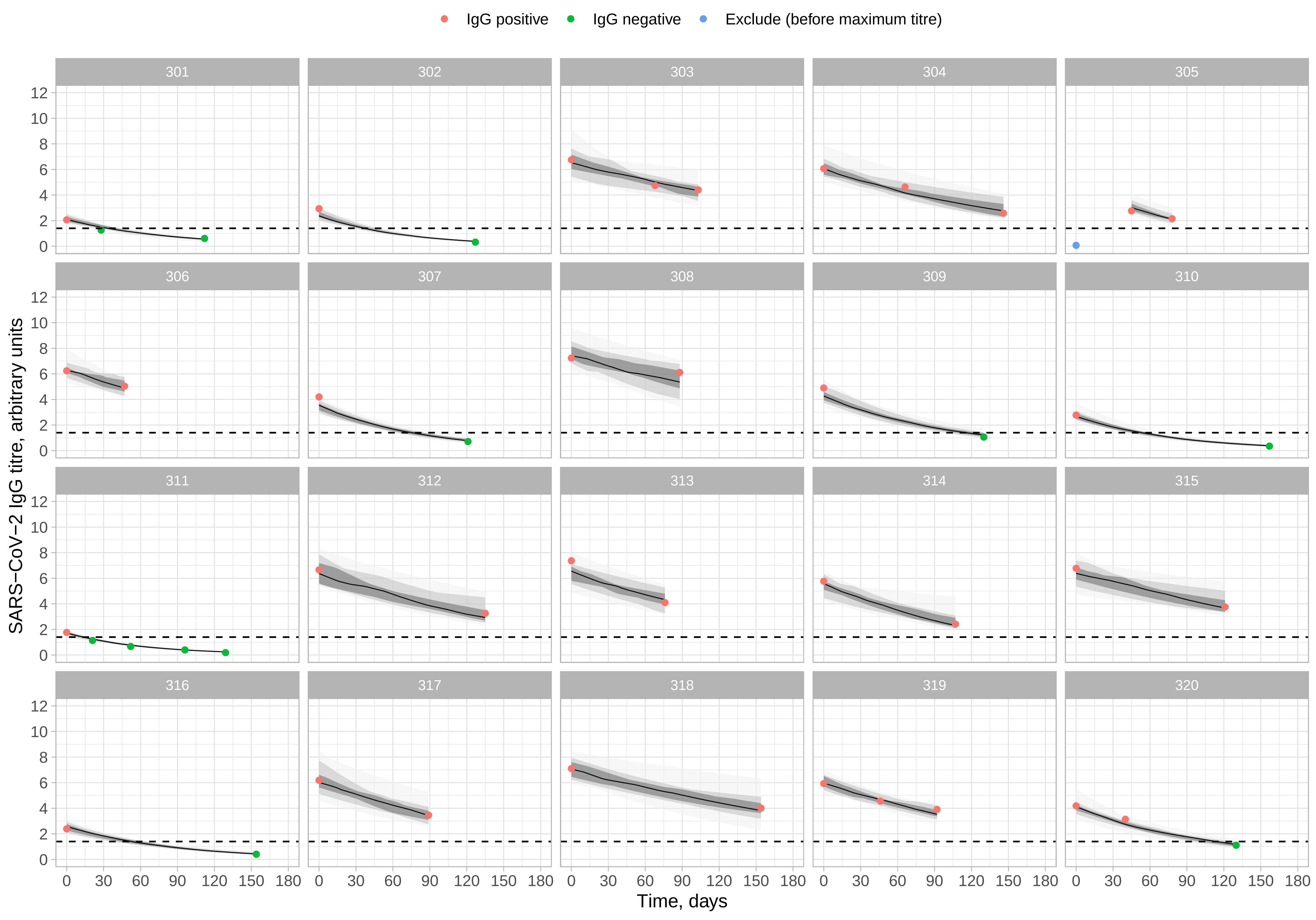

IgG positive
IgG negative
Exclude (before maximum titre)
SARS−CoV−2 IgG titre, arbitrary units
Time, days
0 30 60 90 120 150 180
0 2 4 6 8 10 12

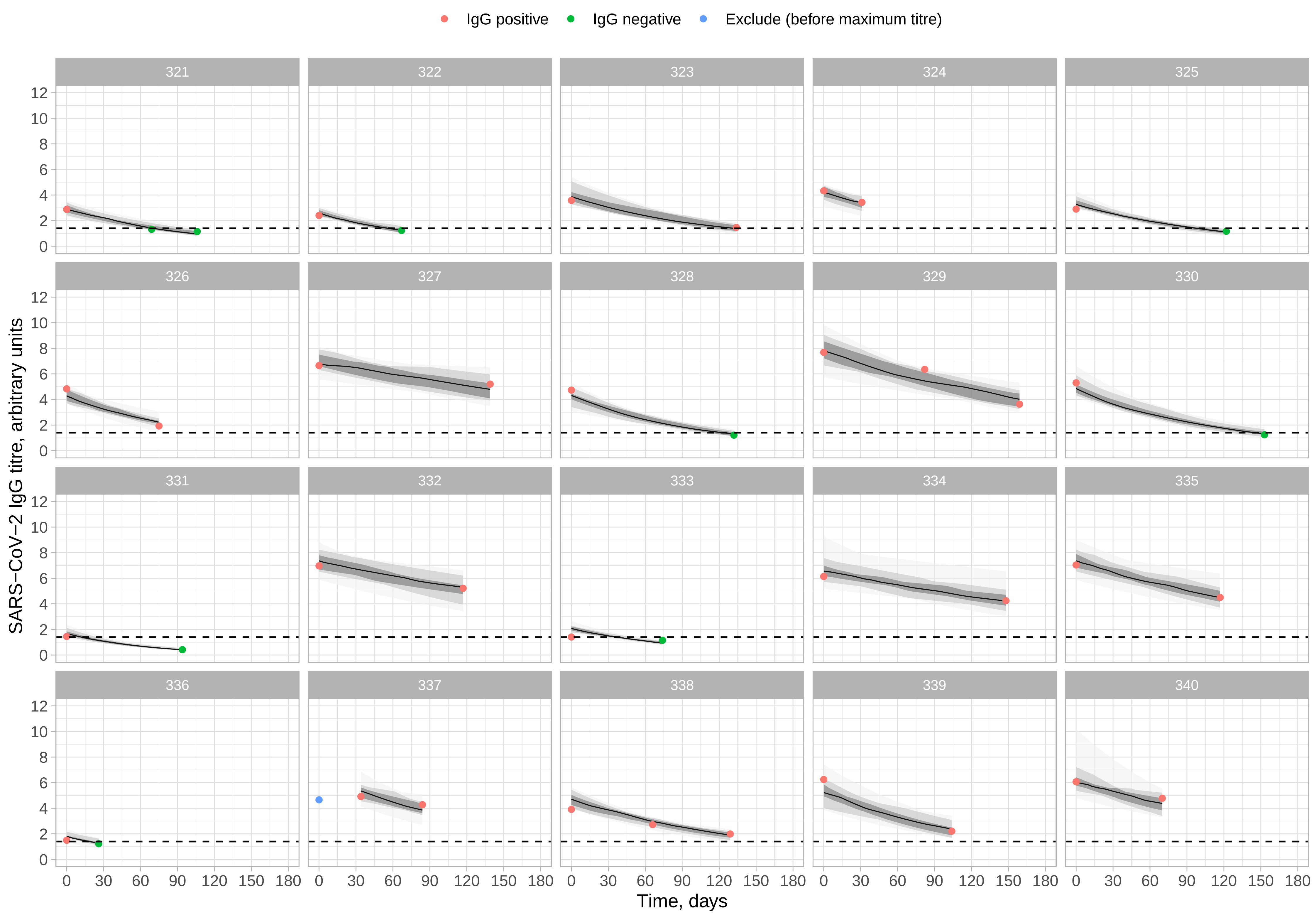

IgG positive
IgG negative
Exclude (before maximum titre)
SARS−CoV−2 IgG titre, arbitrary units
0 2 4 6 8 10 12
0 30 60 90 120 150 180
Time, days

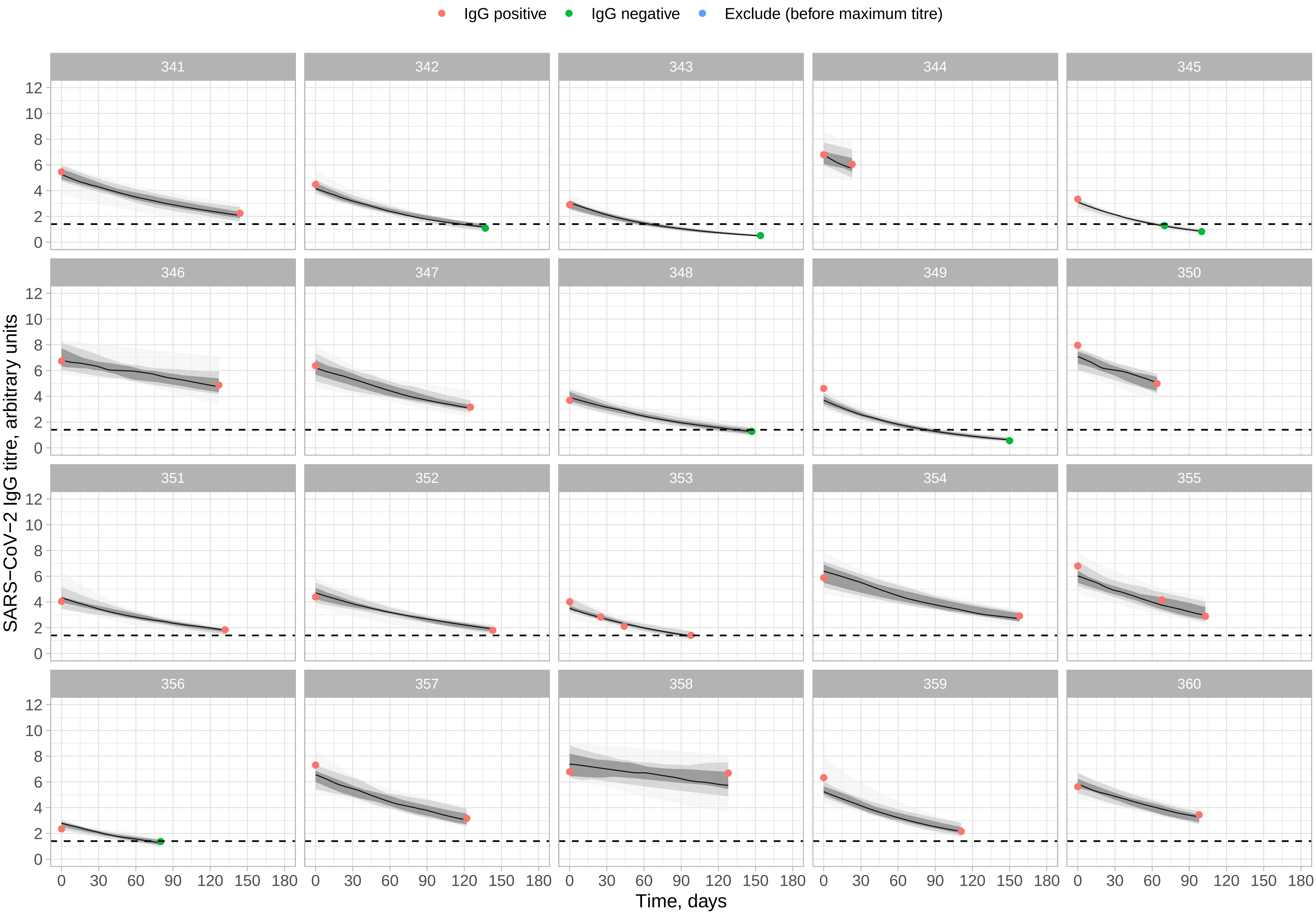

IgG positive
IgG negative
Exclude (before maximum titre)
SARS−CoV−2 IgG titre, arbitrary units
Time, days

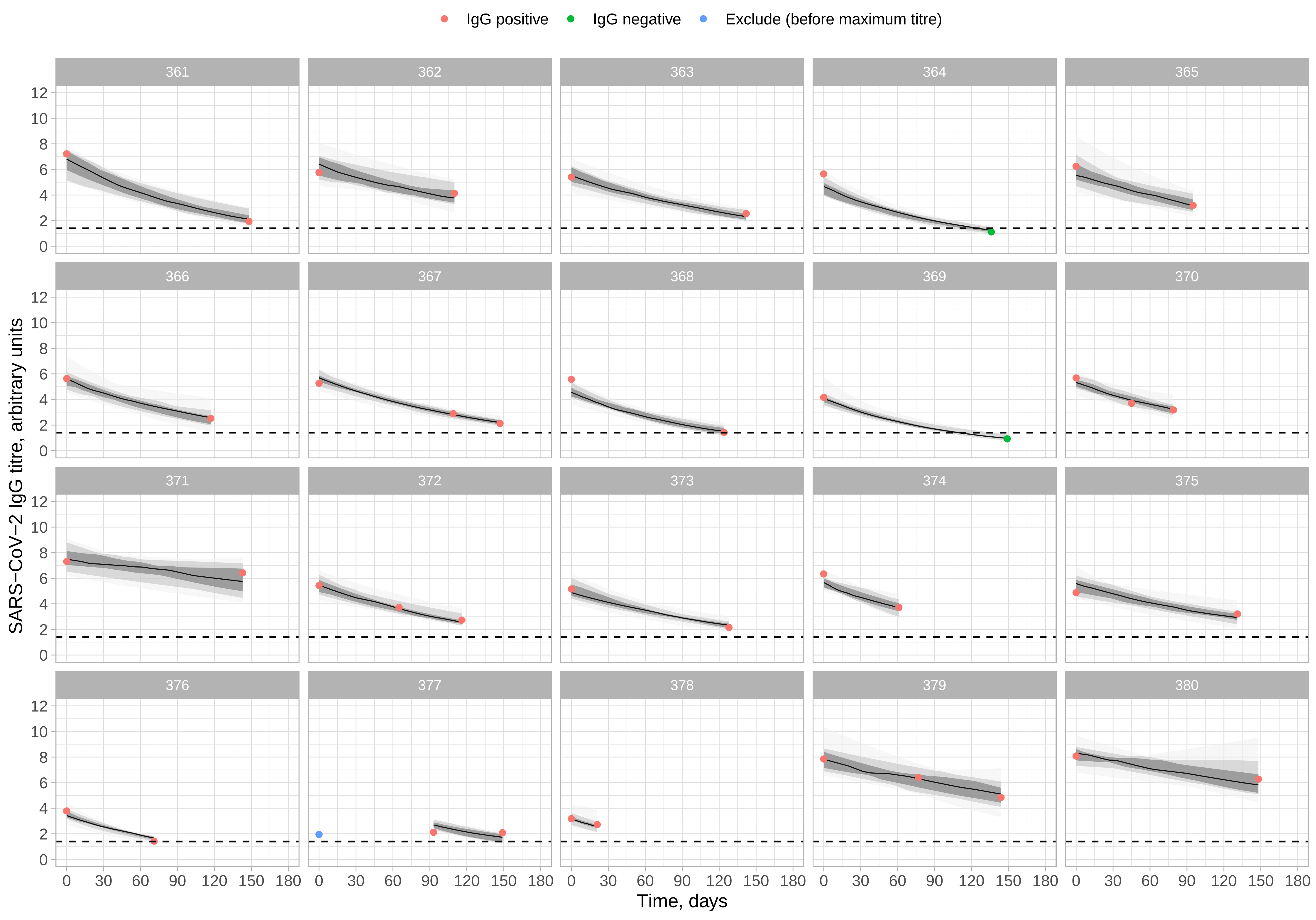
IgG positive
IgG negative
Exclude (before maximum titre)
SARS−CoV−2 IgG titre, arbitrary units
Time, days
0
2
4
6
8
10
12
0
30
60
90
120
150
180

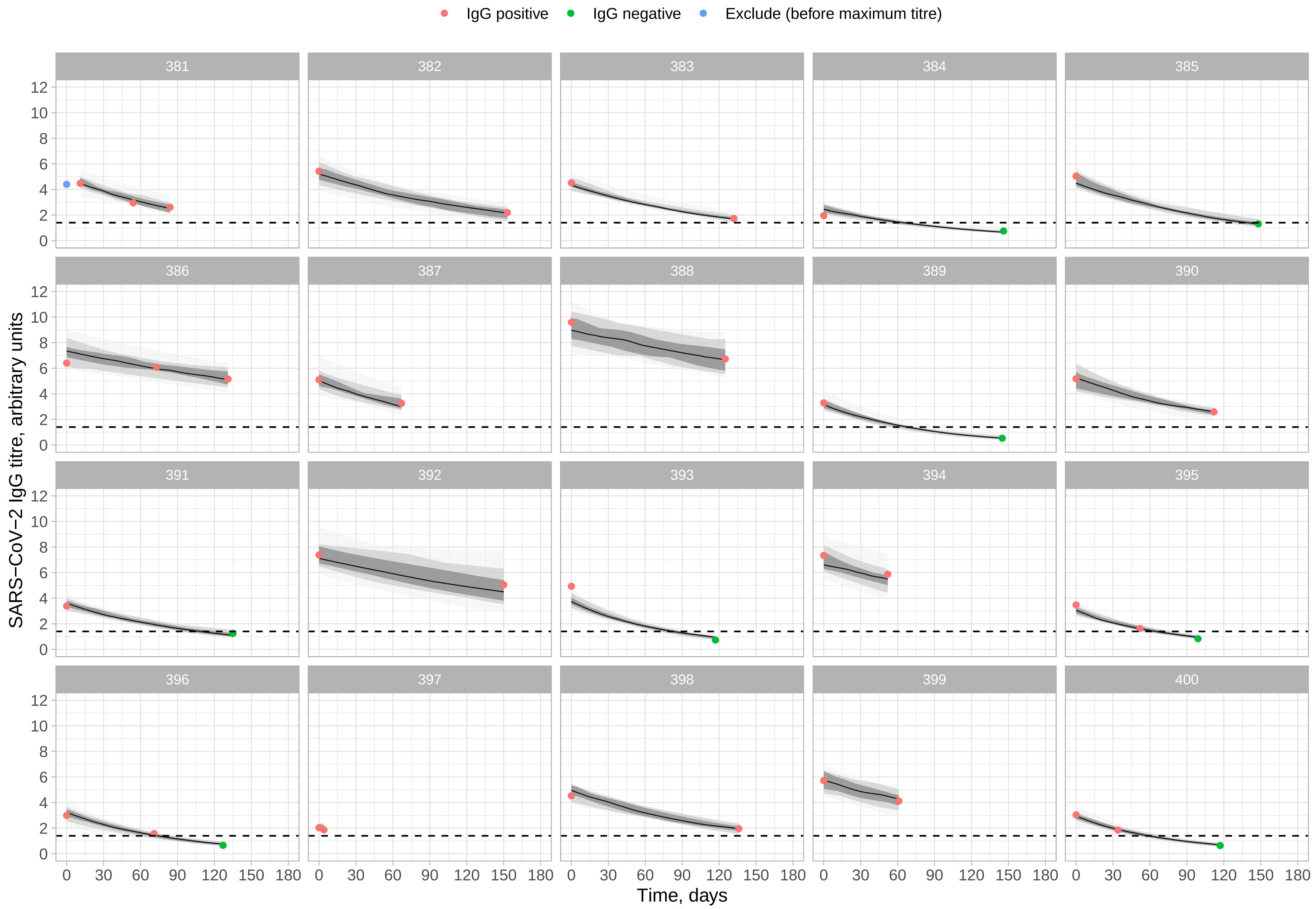

IgG positive
IgG negative
Exclude (before maximum titre)
SARS−CoV−2 IgG titre, arbitrary units
Time, days
0
2
4
6
8
10
12
0
30
60
90
120
150
180

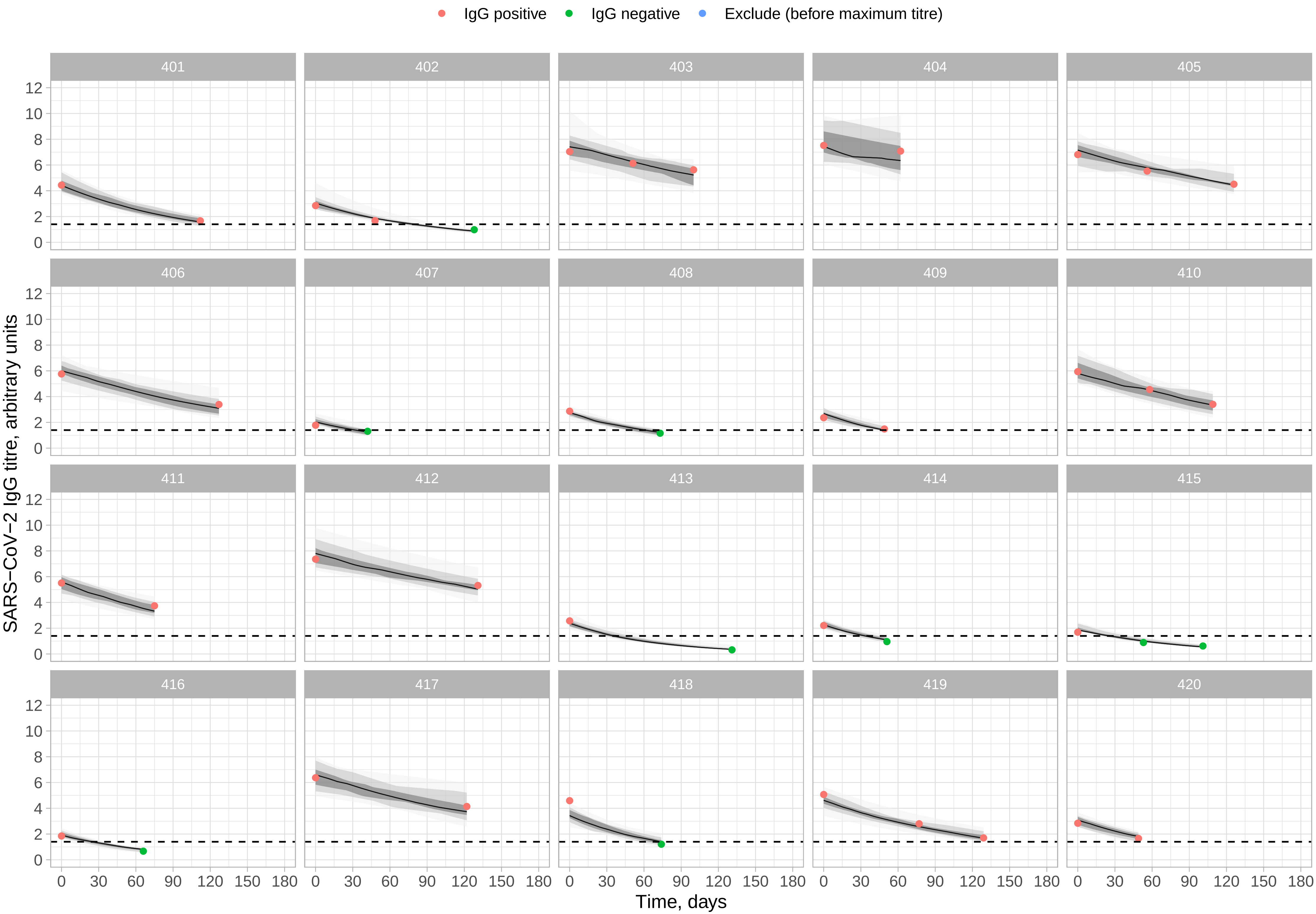
IgG positive
IgG negative
Exclude (before maximum titre)
SARS−CoV−2 IgG titre, arbitrary units
Time, days
0 30 60 90 120 150 180
0 2 4 6 8 10 12

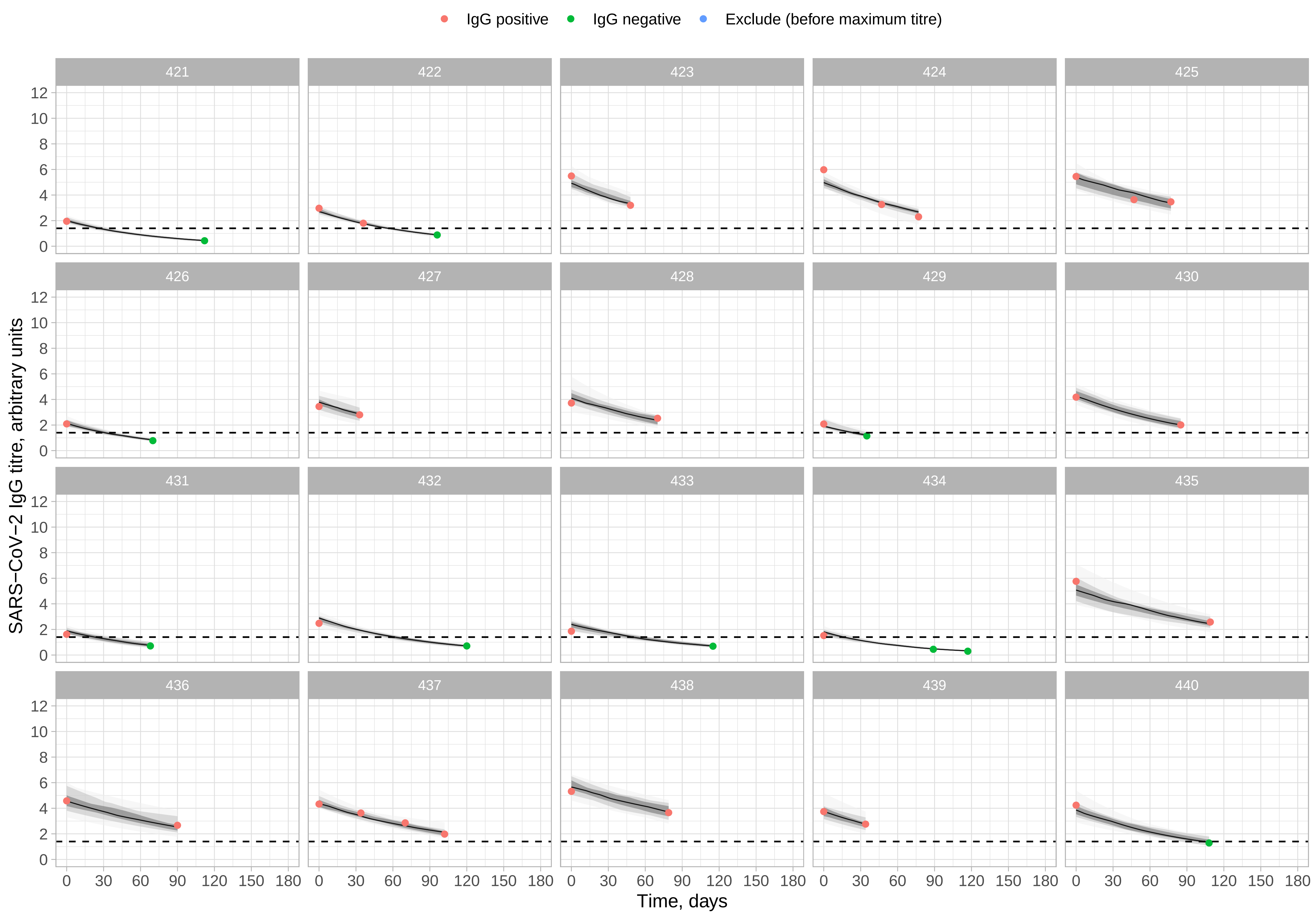
IgG positive
IgG negative
Exclude (before maximum titre)
SARS−CoV−2 IgG titre, arbitrary units
Time, days
0
2
4
6
8
10
12
0
30
60
90
120
150
180

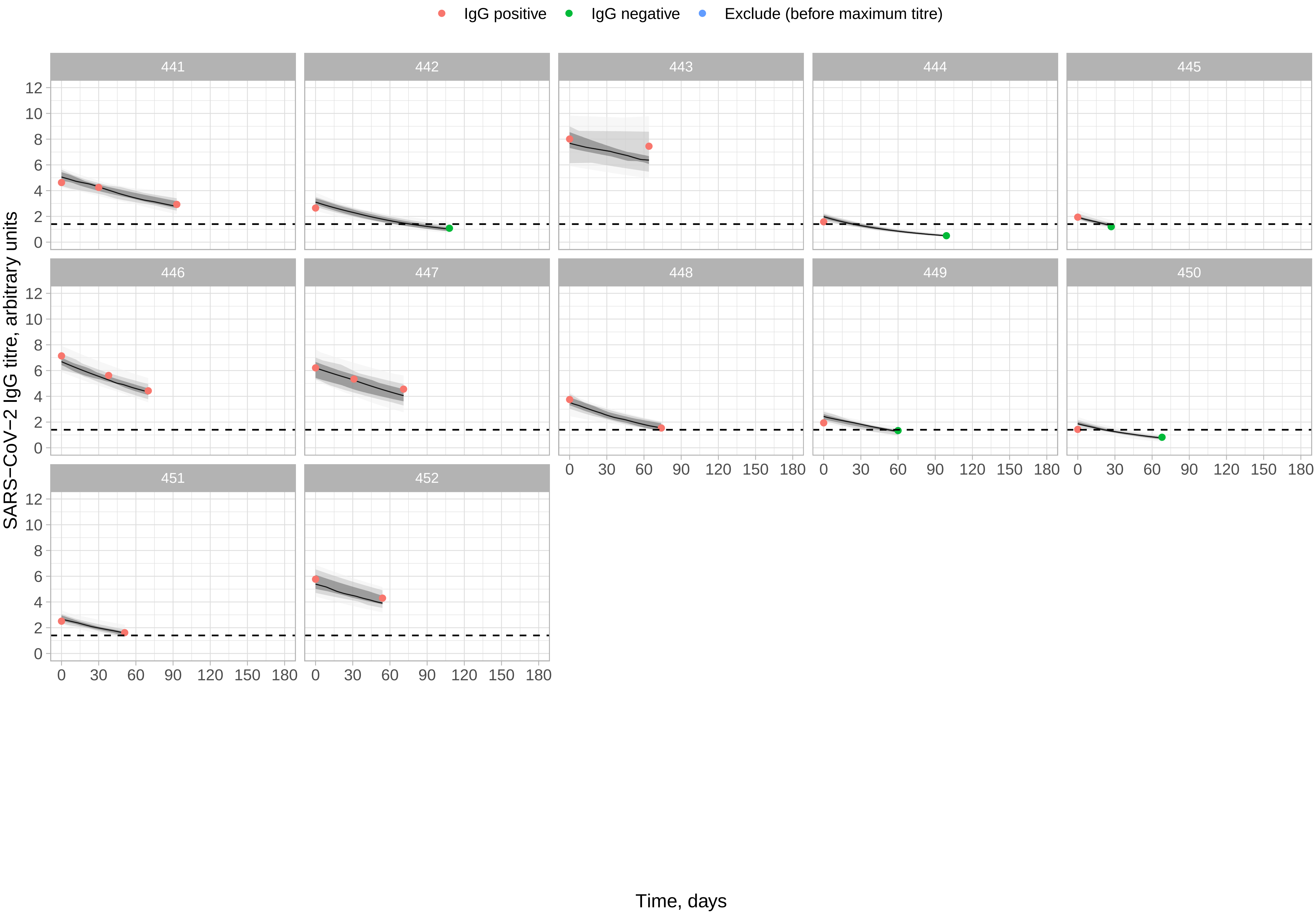

IgG positive
IgG negative
Exclude (before maximum titre)
SARS−CoV−2 IgG titre, arbitrary units
Time, days
0
2
4
6
8
10
12
0
30
60
90
120
150
180
